# Non-coding structural variants identify a commonly affected regulatory region steering *FOXG1* transcription in early neurodevelopment

**DOI:** 10.1101/2025.03.10.25323301

**Authors:** Lisa Hamerlinck, Eva D’haene, Nore Van Loon, Michael B Vaughan, Maria del Rocio Pérez Baca, Sebastian Leimbacher, Lara Colombo, Lies Vantomme, Esperanza Daal, Annelies Dheedene, Himanshu Goel, Björn Menten, Bert Callewaert, Sarah Vergult

## Abstract

The FOXG1 transcription factor is a crucial regulator of embryonic brain development. Pathogenic *FOXG1* variants cause *FOXG1* syndrome. Although structural variants (SVs) in the non-coding region downstream of *FOXG1* have been reported in 38 individuals with similar characteristics, the regulatory pathomechanisms remain unknown.

We identified a *de novo* non-coding deletion in an individual with *FOXG1* syndrome-like disorder, allowing us to delineate a ∼124 kb commonly affected regulatory region (CARR). By integrating epigenomics data, 3D chromatin interaction profiles (Hi-C, UMI-4C), and *in vivo* enhancer assays in zebrafish, we uncovered multiple regulatory elements within this CARR, including a neuronal enhancer cluster and a conserved boundary of the *FOXG1*-containing topologically associating domain (TAD). Hi-C analysis on case lymphoblastoid cells revealed increased interactions with the adjacent TAD. Moreover, sequential UMI-4C and CUT&RUN assays during neural progenitor cell (NPC) differentiation demonstrated dynamic activation of, and interaction with the enhancer cluster. Finally, CRISPR-Cas9 deletion of the enhancer cluster and TAD boundary in NPCs resulted in decreased *FOXG1* transcription.

We identified and characterized enhancer and architectural elements essential for proper *FOXG1* transcription. Our findings provide new insights into chromatin architecture and gene regulation at the *FOXG1* locus, improving SV interpretation in individuals with *FOXG1* syndrome-like disorder.

## INTRODUCTION

The Forkhead Box G1 (FOXG1) transcription factor is highly and specifically expressed in the developing brain, where it acts as an essential transcriptional repressor. It regulates neural progenitor cell proliferation, triggers forebrain patterning, coordinates migration of maturing neurons, and organizes neuronal circuitry assembly^1^. FOXG1 haploinsufficiency has been shown to impair GABAergic interneuron differentiation^2,3^, while FOXG1 overexpression has been associated with overproduction of inhibitory neurons in the context of autism spectrum disorder^4^. Also in the context of schizophrenia, *FOXG1* has been identified as a risk gene by a regulatory, schizophrenia-associated, single-nucleotide polymorphism (SNP)^5^.

Heterozygous aberrations affecting *FOXG1*, leading to FOXG1 loss-of-functio*n*, including intragenic missense, nonsense, frameshift, and even in-frame variants, as well as microdeletions and - duplications, have been associated with a congenital form of Rett syndrome (OMIM #613454), although notable differences with the *MECP2*-associated phenotype have led some to designate it as a distinct disorder, termed *FOXG1* syndrome^1,6–8^. Typical clinical features include severe developmental delay, intellectual disability (ID), microcephaly, hypotonia, seizures and agenesis or hypoplasia of the corpus callosum. In addition, there is a growing cohort of individuals described in literature with structural variants (SVs) disrupting the non-coding region downstream of the *FOXG1* gene^9–24^. Given that the phenotypic characteristics of these individuals closely resemble those associated with *FOXG1* syndrome, we refer to this condition as *FOXG1* syndrome-like disorder from now on. Although the putative regulatory effects have not been functionally characterized, it has been postulated that these copy-number variants (CNVs; deletions, duplications) as well as balanced chromosomal aberrations (BCAs; translocations, inversions) 3’ to *FOXG1* disrupt regulatory mechanisms governing *FOXG1* transcription.

These cases underline that FOXG1 expression must be tightly regulated to support normal brain development in agreement with its established dosage sensitivity^3^, but little is known about the gene regulatory elements controlling *FOXG1* transcription. The gene-poor region downstream of *FOXG1* contains several *in vivo* validated enhancer elements displaying activity in neural tissues^25^, yet their effect on FOXG1 expression has not been investigated. Additionally, while the 3D chromatin conformation of the locus, including its organization in topologically associating domains (TADs), has been hypothesized to be crucial for regulating FOXG1 expression^10,26^, the impact of TAD disruption on *FOXG1* regulation and transcription remains unexplored.

In this study, we present an overview of all previously reported individuals (n=38)^10–24^ with an SV overlapping the *FOXG1* TAD. We report a first individual with a non-coding microdeletion 3’ to *FOXG1* allowing to greatly narrow down the previously identified smallest region of overlap (∼400kb)^10^ to a non-coding commonly affected regulatory region (CARR) of ∼124kb. Using chromatin interaction mapping via UMI-4C and *in vivo* enhancer assays in zebrafish, we show that the *FOXG1* promoter interacts with this region during human neuronal development and that the CARR contains a neuronal enhancer cluster (EC) and a TAD boundary. Given the crucial role of FOXG1 during NPC proliferation and differentiation, we perform a series of CUT&RUN assays (H3K4me3, H3K4me1, H3K27ac) during NPC differentiation to elucidate temporal enhancer dynamics in the *FOXG1* locus. Furthermore, we create *in vitro* knock-out models using CRISPR-Cas9 to assess the impact of both EC and TAD boundary deletion on *FOXG1* transcription during NPC differentiation.

In summary, we narrowed down a CARR downstream of *FOXG1,* which is disrupted in a cohort of individuals with *FOXG1* syndrome-like characteristics, and contains both enhancer and TAD boundary elements. Our results greatly improve the functional annotation and validation of regulatory elements at the *FOXG1* locus, which is crucial for correct SV interpretation.

## RESULTS

### Non-coding deletion in individual with FOXG1 syndrome-like disorder narrows down commonly affected regulatory region 3’ to FOXG1

To gather a comprehensive overview of reported individuals with a *FOXG1*-related phenotype in which the *FOXG1* gene itself is intact, we mined literature and DECIPHER (DatabasE of genomiC varIation and Phenotype in Humans using Ensembl Resources) for SVs within or overlapping the *FOXG1* TAD, not affecting the *FOXG1* coding sequence. (**Fig 1a, Sup Table S1**). We found 20 individuals with CNVs (1 duplication, 19 deletions), ranging in size from 391 kb to almost 6 Mb, 17 cases with BCA breakpoints downstream of *FOXG1*, as well as one individual with a ring chromosome resulting from a ~50Mb interstitial excision, all displaying phenotypic overlap with *FOXG1* syndrome-like features. While most of these SVs occurred *de novo* (31/39), the duplication (dup1) was maternally inherited. For five deletions (del1, 2, 5, 7 & 16) and 2 BCAs (bca11 & 17), the inheritance pattern is unknown. Although we found an additional three CNVs (1 deletion and 2 duplications) overlapping the *FOXG1* TAD without affecting the *FOXG1* coding sequence, we were unable to obtain any phenotypic information, and therefore were excluded from the overview (ex.del1; ex.dup1&2) (**Sup Table S1, Sup Fig S2**).

**Fig 1.**
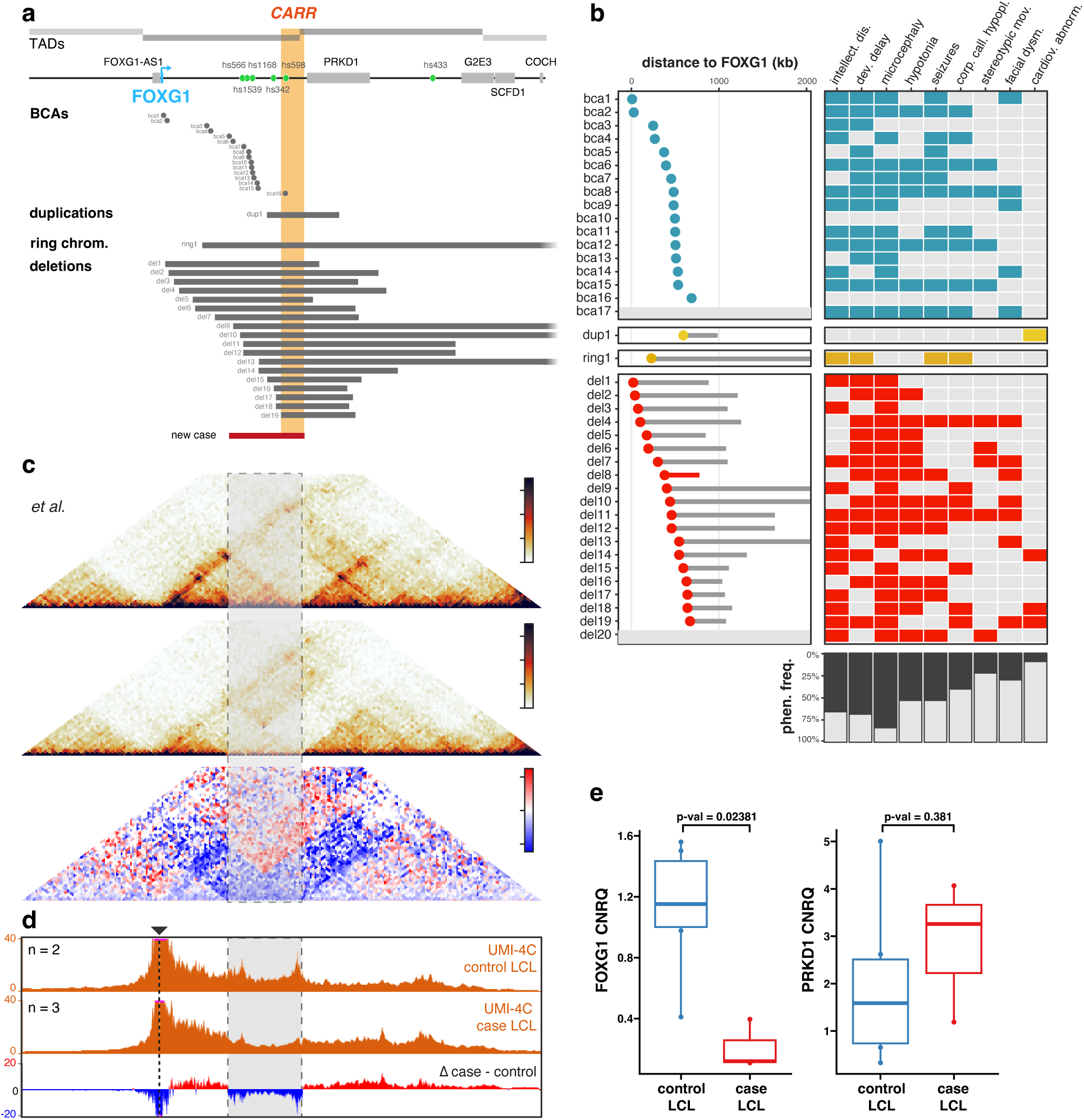
Structural variation disrupts the *FOXG1* regulatory region in individuals with *FOXG1* syndrome-like characteristics. (a) Schematic view of structural variants (SVs) located 3’ to *FOXG1*, obtained from literature, DECIPHER (grey) and the in-house identified case (red). (b) Schematic overview of each SV annotated with its distance to the *FOXG1* transcription start site, along with associated phenotypic outcomes. These include neurodevelopmental abnormalities (intellectual disability, developmental delay, microcephaly, hypotonia, seizures, corpus callosum hypoplasia, stereotypic movements and facial dysmorphisms) primarily linked to aberrant *FOXG1* transcription and cardiovascular abnormalities possibly related to aberrant *PRKD1* transcription. (c-d) Differential 3D chromatin interactions at the *FOXG1* locus between case and control LCLs using Hi-C (c), and UMI-4C (d). (e) Differential RNA expression between six independent control LCLs and case LCLs for *FOXG1* (Mann-Whitney U test, p = 0.02381) and *PRKD1* (Mann-Whitney U test, p = 0.381).

We employed POSTRE^27^, an *in silico* prediction tool, for an initial assessment of the pathogenicity and underlying pathomechanisms of these SVs. For each candidate gene within the affected TADs, independent pathogenic prediction scores (PS) were made using cell-type or tissue-specific genomic data, particularly neurodevelopmental datasets (pfcGw15 and pfcGw18), including gene expression profiles, enhancer and TAD maps. In all cases *FOXG1* was considered the most likely causal gene (causative gene PS>0.8, **Sup Table S2**). This aligns with the phenotypic data we gathered, as almost all were reported to display hallmarks of *FOXG1* syndrome, with the most frequently reported symptoms including ID, developmental delay, microcephaly, hypotonia, seizures, stereotypic movements and hypoplasia of the corpus callosum (**Fig 1b, Sup Table S1**). However, the duplication included in the overview remains difficult to definitively link to a *FOXG1*-related disease mechanism, as it was maternally inherited and the phenotype does not align with the other *FOXG1* syndrome-like cases.

Here, we also report a novel case (del8) with a *de novo* ∼416kb deletion downstream of *FOXG1* (finemapped to chr14:29,143,605-29,559,423 (hg38)) (**Fig 1a, Sup Fig S1).** Clinical characteristics indicated a *FOXG1*-related disease mechanism and include severely delayed neuromotor development, epileptic seizures, microcephaly, chorea, and hypotonia (**Sup Text** for full clinical description). Based on the distal breakpoint in this individual, we could narrow down a ∼124 kb commonly affected regulatory region (CARR) situated at chr14:29,435,515-29,559,423 (hg38) (**Fig 1a, Sup Fig S1**). This greatly narrows down the previously reported smallest region of overlap, which spanned ∼400kb^10^. Of note, this individual (del8) represents the first case with a CNV downstream of *FOXG1* that does not overlap the *PRKD1* gene (**Fig 1a, Sup Fig S1**), thereby excluding any involvement of direct *PRKD1* aberrations in the phenotype. Heterozygous *de novo* missense mutations in *PRKD1* have been identified in individuals with syndromic congenital heart defects (pulmonic stenosis, septal defects), with additional clinical features including developmental delay, ectodermal and limb abnormalities (OMIM #617364)^28,29^. Intriguingly, although all 20 previously reported CNVs downstream of *FOXG1* also affect *PRKD1*, cardiac defects were only reported in four cases (del14, del18, del19, dup1) (**Fig 1a-b, Sup Table S1**). The previously reported missense mutations in PRKD1 were predicted to result in both gain- and loss-of-function, although clinical characteristics associated with both types of mutations are similar^28,29^ Yet, the absence of cardiac defects in most cases with CNVs overlapping *PRKD1* suggests that PRKD1 loss (due to CNVs) and PRKD1 (in)activation (due to missense mutations) have different phenotypic consequences.

### Non-coding deletion affects 3D chromatin interactions at the FOXG1 locus

Since non-coding SVs can have an impact on gene regulation, 3D chromatin structure, and ultimately can influence gene expression, we assessed the impact of the 416 kb non-coding deletion identified in the new *FOXG1* syndrome-like case (del8) on *FOXG1* regulatory interactions, TAD structure, and transcription. We therefore performed UMI-4C^30^ and *in situ* Hi-C^31^ on a lymphoblastoid cell line (LCL) from this individual and compared interaction profiles/matrices to those from a control LCL (control LCL Hi-C data previously generated by Melo *et al.*^13^). Both Hi-C matrices and UMI-4C profiles from case LCL indicated removal of the distal *FOXG1* TAD boundary and increased interactions with the adjacent TAD (**Fig 1c-d**). In addition to these structural changes, mRNA expression analysis revealed a significant decrease in *FOXG1* transcription in the case’s LCL compared to six independent control LCLs (**Fig 1e**; Mann-Whitney U test, p = 0.02381). Notably, the deletion did not directly affect the transcription of the neighbouring *PRKD1* gene, regardless of the observed fusion of the *FOXG1* and *PRKD1* TADs (**Fig 1e**; Mann-Whitney U test, p = 0.381).

### Commonly affected regulatory region harbours enhancer cluster

The phenotypic overlap of the individuals with *FOXG1* syndrome-like characteristics suggests that the CARR may play a key role in regulating *FOXG1* transcription during neurodevelopment. To determine whether this region could indeed be involved in *FOXG1* regulation, we mapped chromatin interactions using UMI-4C^34^ for the *FOXG1* promoter in induced pluripotent stem cells (iPSCs), neural stem cells (NSCs), neural progenitor cells (NPCs) and neurons. UMI-4C profiles indicated frequent chromatin looping between the *FOXG1* promoter and several previously validated VISTA enhancers as well as the CARR, in concordance with previously published Hi-C data from human fetal cortex (**Fig 2c**)^32^. Interestingly, interaction frequencies varied between cell types, with many interactions becoming more prominent in NPCs and neurons compared to NSCs and iPSCs.

**Fig 2.**
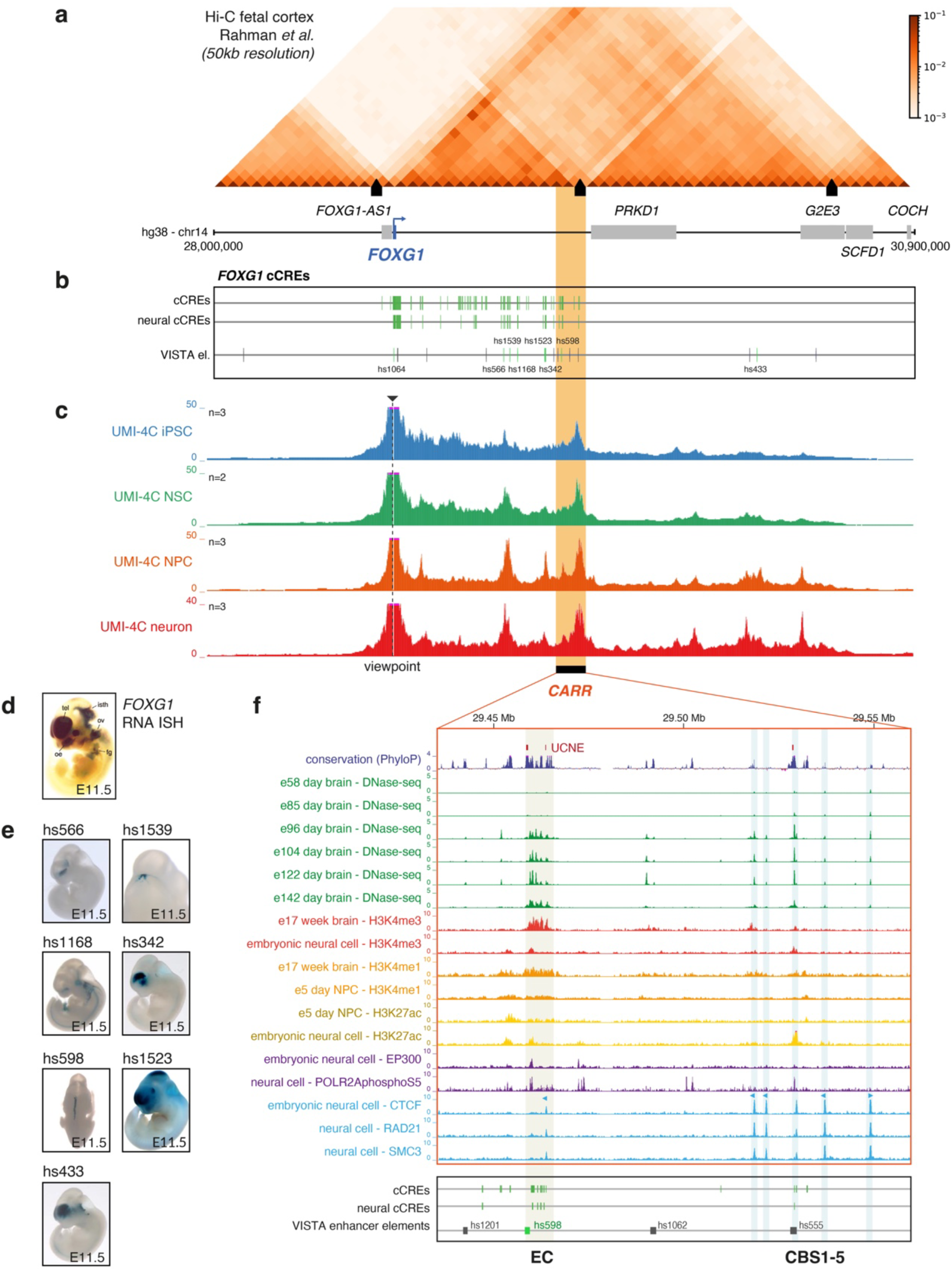
*FOXG1* chromatin interactions and regulatory landscape. (a) TAD structure of the *FOXG1* locus as shown by the Hi-C interaction frequency matrix (fetal cortex, 50kb resolution, Rahman *et al*.32). TAD boundaries are indicated by black boxes. (b) Candidate *cis*-regulatory elements (cCREs) within the *FOXG1* locus. Identified (neural) enhancer-like cCREs based on human DNase I hypersensitive sites (DHSs)33 and ENCODE SCREEN34 enhancer-like elements, as well as validated VISTA enhancer elements (positive: green, negative: grey)25. Commonly affected regulatory region (CARR) indicated in orange (chr14:29,435,515-29,559,423, hg38). (c) UMI-4C interaction frequency profiles (normalized and smoothed) with the *FOXG1* promoter as viewpoint (dashed line) in induced pluripotent stem cells (iPSCs, n=3), neural stem cells (NSCs, n=2), neural progenitor cells (NPCs, n=3) and neurons (n=3). (d) LacZ staining for Cre recombination in Foxg1-Cre mice illustrates Foxg1 expression patterns in E11.5 mouse embryos (telencephalon (tel), mid-hind brain region (isthmus, isth), olfactory epithelium (oe), otic vesicle (ov), lens, retina, foregut (fg))36. (e) Activity patterns of validated enhancer elements from the VISTA enhancer browser in E11.5 mouse embryos (hs566: forebrain; hs1539: hindbrain; hs1168: cranial nerve, facial mesenchyme, hindbrain; hs342: forebrain; hs598: neural tube; hs1523: branchial arch, eye, facial mesenchyme, forebrain, midbrain, hindbrain; hs433: forebrain, hindbrain)25. (f) Closeup of CARR, showing PhyloP basewise conservation with ultra-conserved non-coding elements (UCNEs)35, accessible chromatin through DNase-seq in embryonic brain, ChIP-seq for regulatory markers H3K4me3, H3K4me1, H3K27ac, as well as active RNA polymerase II (POLR2AphosphoS5) and the P300 cofactor in (embryonic) neural cells and NPCs, ChIP-seq for the architectural protein CTCF and cohesin subunits RAD21 and SMC3 in (embryonic) neural cells, (neural) cCREs, validated VISTA enhancer elements (positive: green, negative: grey)25. Highlights show regions of interest: enhancer cluster (EC) and CTCF-binding site (CBS) 1-5.

Next, we sought to assess the regulatory potential of the CARR. Despite the presence of multiple validated brain enhancer elements 3’ to *FOXG1*, the only validated enhancer within the CARR is element hs598 (**Fig 2b**)^25^. Given that its activity pattern appears to be restricted to the neural tube, this enhancer element likely contributes only partially to establishing the full FOXG1 expression pattern (**Fig 2d-e**). Deletion of this single element is therefore unlikely to be sufficient to disrupt *FOXG1* transcription to such an extent as to result in the disease phenotype. Therefore, we compiled a list of candidate *cis*-regulatory elements (cCREs) present within the *FOXG1* TAD based on tissue-wide maps of human DNase I hypersensitive sites (DHSs) generated by Meuleman *et al.*^33^ Specifically, we cross-referenced neural DHS maps with ENCODE’s SCREEN^34^ registry of candidate regulatory elements to identify all neural DHSs with an enhancer-like epigenomic signature. This resulted in the identification of 99 candidate neural enhancers within the *FOXG1* TAD (**Fig 2b**), of which eight were located within the CARR (**Fig 2f**). Visualization of publicly available epigenomic datasets from ENCODE^34^, including DNase-seq data from different stages of human embryonic brain development and ChIP-seq for several regulatory markers, indicated that five of the neural cCREs cluster together in a conserved region with strong regulatory potential, which we termed the enhancer cluster (EC) region (chr14:29458444-29465794, hg38; **Fig 2f, Sup Fig S3**). Notably, the EC region also included the previously validated hs598 enhancer, which contains an ultraconserved non-coding element (UCNE)^35^. However it neither overlaps with the strongest epigenetic signals within the EC region, nor any of the neural cCREs, indicating that additional, untested sequences within the EC might have strong enhancer potential (**Fig 2f, Sup Fig S3**).

### Commonly affected regulatory region plays a role in establishing TAD structure at the FOXG1 locus

In addition to the presence of a putative enhancer cluster, there are several indications that the CARR plays an important role in establishing 3D chromatin structure at the *FOXG1* locus. Inspection of Hi-C data from human fetal cortex and UMI-4C interaction data for the *FOXG1* and *PRKD1* promoters suggested that the *FOXG1* gene is located near the centromeric boundary of a megabase-scale TAD, containing two distinct subdomains (**Fig 2a, Sup Fig S4**). Insulation score analysis on the Hi-C matrix indeed indicated the presence of a TAD boundary within the CARR. Consulting published Hi-C datasets from different human tissues and cell types, including embryonic stem cells, neural progenitor cells and prefrontal cortex, the TAD structure of the *FOXG1* locus appeared to be largely invariant (**Fig 2c, Sup Fig S5**).

In addition, ChIP-seq data for binding of the architectural protein CTCF, as well as cohesion subunits RAD21 and SMC3, indicated the presence of a cluster of five CTCF-binding sites (CBS1-5) within the CARR, constituting the *FOXG1* TAD boundary (**Fig 2f, Sup Fig S6,S7)**. CTCF motifs at three out of five CBSs are convergently orientated to the centromeric TAD boundary proximal to *FOXG1*, indicating these CBSs are likely involved in establishing interactions within the *FOXG1* TAD through a loop extrusion mechanism. CTCF binding at these sites appeared to be largely invariant across tissues and cell types, in concordance with the conserved 3D genome structure of the locus (**Sup Fig S6)**. Interestingly, the presence of enhancer marks and ultraconserved sequences at CBS3 might be indicative of an underlying enhancer element (**Fig 2f, Sup Fig S3**). This element (hs555) lacked enhancer activity in *in vivo* enhancer assays^25^. However, Won *et al.* found that a 500bp sequence surrounding a schizophrenia-associated SNP (rs1191551) approximately 1.5kb downstream of CBS3 showed enhancer activity in luciferase assays and its deletion decreased *FOXG1* expression by 20-30% in HEK293 cells^5^. This suggests that enhancer potential at the CBS region cannot be fully excluded yet.

### Candidate enhancers display activity in neural tissues during zebrafish development

To assess the tissue-specific enhancer activity of cCREs within the EC region, we delineated five likely candidate regions (cCREa-cCREe) based on conserved sequence stretches and epigenetic signatures in human brain samples (**Fig 3a**). These sequences were subsequently cloned and tested through two independent *in vivo* zebrafish enhancer assays (i.e., E1b vector^37^ and ZED vector^38^) and GFP expression was monitored at 24, 48, and 72 hours post-fertilization (hpf) (**Fig 3b, Table S3**). cCREa was chosen to be largely similar to VISTA element hs598 and, in concordance with the results from the VISTA enhancer browser, also showed consistent GFP expression within the zebrafish notochord. Furthermore, cCREa drove GFP expression in the forebrain, otic vesicle, and spinal cord in the ZED vector assay (**Fig 3b**). cCREb also displayed consistent activity in the notochord and drove GFP expression in the forebrain in the ZED vector assay (**Fig 3b**). cCREc-driven GFP expression was observed in the forebrain and olfactory pit (**Fig 3b**). GFP expression driven by cCREd was consistently localized in the otic vesicle. In the ZED vector assay, cCREd also drove GFP expression in the forebrain and in the heart (**Fig 3b**). Enhancer activity in the heart was also observed for cCREe in the E1b vector assay, while the ZED vector assay revealed activity in the forebrain for this element (**Fig 3b**). In sum, all five identified enhancer elements exhibited consistent enhancer activity in concordance with previously described *Foxg1* expression patterns^39–44^ and may thus regulate *FOXG1* expression. An overview of all percentages of alive embryos with observed GFP expression at 24, 48 and 72 hpf can be found in **Sup Table S3 and Sup Fig S8**.

**Fig 3.**
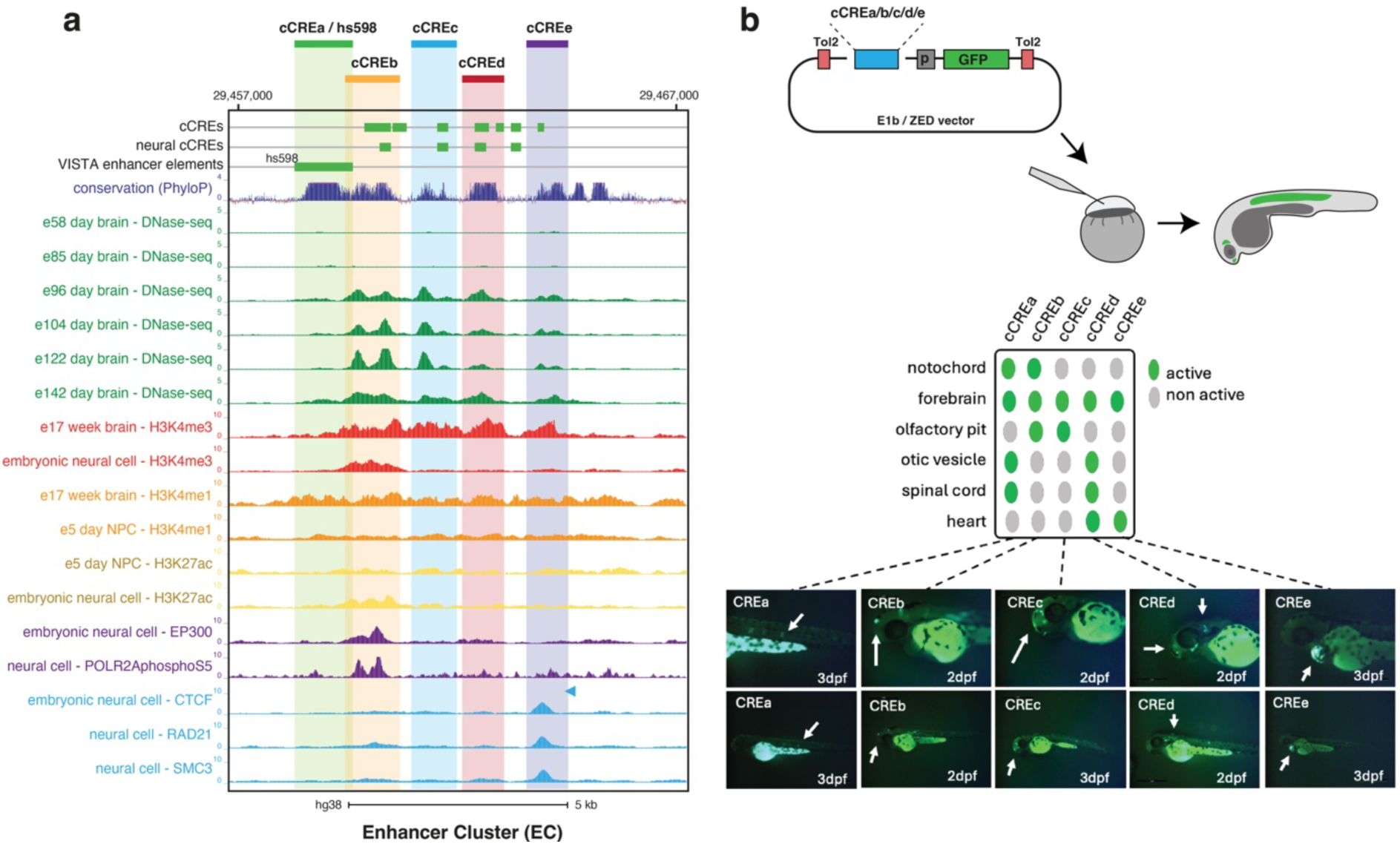
Candidate *cis*-regulatory elements (cCREs) show tissue specific enhancer activity. (a) Close-up of the five tested candidate *cis*-regulatory elements (cCREs) (cCREa-cCREe) identified based on conserved sequence stretches and publicly available epigenetic signatures in human fetal brain samples (b) Summary of *in vivo* enhancer assays in zebrafish F0 transgenic lines using both the ZED38 and E1B37 vector; box indicating in which tissues GFP reporter expression was observed (notochord, forebrain, olfactory pit, otic vesicle, spinal cord, and heart); representative image of reporter expression at 48 or 72 hours post-fertilization (hpf) for each cCRE (white arrows). cCREa: notochord; CREb: forebrain; cCREc: forebrain and olfactory pit; cCREd: otic vesicle (vertical) and forebrain (horizontal); cCREe: heart.

### FOXG1 is dynamically regulated during the early phases of NPC differentiation

FOXG1 expression is a well-known marker of forebrain NPCs^39^ and reanalysis of publicly available single-cell RNA-seq data from human brain development indeed mainly showed high *FOXG1* expression in ventral telencephalic NPCs and neuron^45^ (**Sup Fig S9**). Given FOXG1’s established role in NPC proliferation and differentiation, we investigated the role of the EC and other regulatory elements in controlling *FOXG1* expression during NPC differentiation. First, to evaluate the expression of *FOXG1* and other marker genes, we differentiated iPSCs into NPCs, using four wildtype cell lines, over a 12-day differentiation experiment. Although we noted variability in *FOXG1* expression between the different biological replicates, reproducible peak *FOXG1* expression was observed between days 6 and 8 of NPC differentiation (**Fig 4a**). To investigate whether the variability of *FOXG1* expression was attributable to a heterogeneous NPC population, we assessed the expression of various NPC markers for different brain regions: *FOXG1*, *DLX2*, *SIX3*, and *OTX2* (forebrain); *PAX6* (forebrain/midbrain); *LMX1A* (midbrain); *EN1* and *IRX3* (midbrain/hindbrain); and *HOXB4* (hindbrain/spinal cord) (**Sup Fig S10**). Despite the lack of a clear correlation among most of these markers, the observed variability in expression across other NPC markers (**Sup Fig S10**) suggests that differences in the NPC population might contribute to the observed variability in *FOXG1* expression.

**Fig 4.**
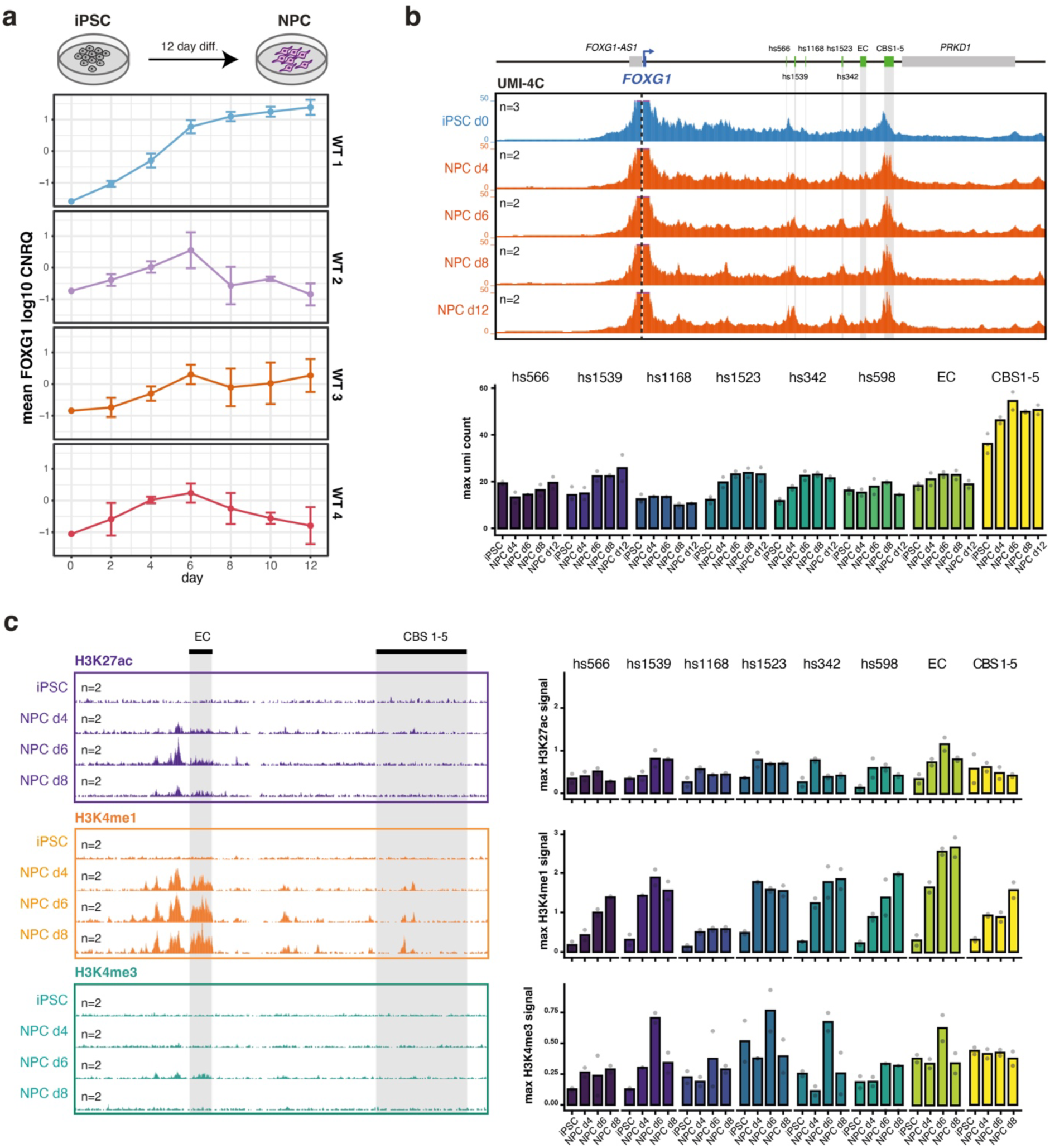
*FOXG1* regulatory dynamics during neural progenitor cells (NPC) differentiation. (a) Schematic overview of the iPSC-derived NPC differentiation. RNA expression of *FOXG1* during a 12-day NPC differentiation for four biological wildtype (WT) replicates. (b) *FOXG1* promoter interaction frequencies using UMI-4C, during a 12-day NPC differentiation. Interactions with validated VISTA enhancer elements and the enhancer cluster (EC) are highlighted in grey. (c) Characterization of the *FOXG1* cis-regulatory landscape in iPSC-derived NPCs. (left) CUT&RUN tracks for H3K27ac, H3K4me1 (active enhancer marks) and H3K4me3 (active promoter mark) surrounding the commonly affected regulatory region (CARR) within the *FOXG1* TAD, including the EC and TAD boundary (grey), during an 8-day NPC differentiation. (right) Maximal (max) signal observed for H3K27ac, H3K4me1 and H3K4me3 over an 8-day NPC differentiation at validated VISTA enhancer elements, the EC and TAD boundary.

### The enhancer cluster is activated during early NPC differentiation and shows increased contact with FOXG1

To understand the regulatory mechanisms underlying FOXG1 induction during NPC differentiation, we characterized the epigenetic landscape of active enhancers and promoters. CUT&RUN analysis revealed enrichment of H3K27ac and H3K4me1, active enhancer marks, at the EC region upon NPC differentiation (**Fig 4c**). Interestingly, we also observed dynamic enrichment of these active enhancer marks at an adjacent region upstream of the EC (**Fig 4c**). Furthermore, we examined active enhancer marks at the previously validated VISTA enhancer elements (**Fig. 4c**). These elements exhibited lower overall signal, with a prominent peak observed only at day 6 of NPC differentiation. Additionally, H3K4me3 and H3K27ac, active promoter marks, were observed at the promoters of several NPC marker genes (**Sup Fig S11**), indicating successful differentiation. UMI-4C experiments further demonstrated increased chromatin interaction frequency between the EC and the *FOXG1* promoter during differentiation (**Fig 4b**), suggesting a regulatory role for the elements within the EC in controlling *FOXG1* expression. Further analysis of VISTA enhancer elements revealed a similar dynamic interaction with the *FOXG1* promoter during NPC differentiation. The interactions between the *FOXG1* promoter and the enhancer elements overall reach their peak around days 6-8 of differentiation, consistent with the activation of the enhancers and *FOXG1* transcription.

### Deletion of functional elements in the commonly affected regulatory region results in decreased FOXG1 expression

Next, we assessed the impact of removing the regulatory elements within the CARR, *i.e.* the EC and TAD boundary, on *FOXG1* expression in the NPC differentiation model characterized above. We generated heterozygous *in vitro* knockout (KO) models targeting the EC region and the TAD boundary through CRISPR-Cas9 in iPSCs, using paired guide RNAs (gRNAs) complementary to both sides of the targeted regions (**Fig 5a**). Upon single-cell isolation and deletion screening, we selected edited clones and matched controls (i.e. wildtype clones that underwent the same experimental procedure). Subsequently, we performed three independent experiments in which we differentiated up to six biological replicates per genotype (control: 6 clones, EC DEL^HET^: 3 clones, TAD DEL^HET^: 6 clones) from iPSCs to NPCs over the course of eight days (**Fig 5a**).

**Fig 5.**
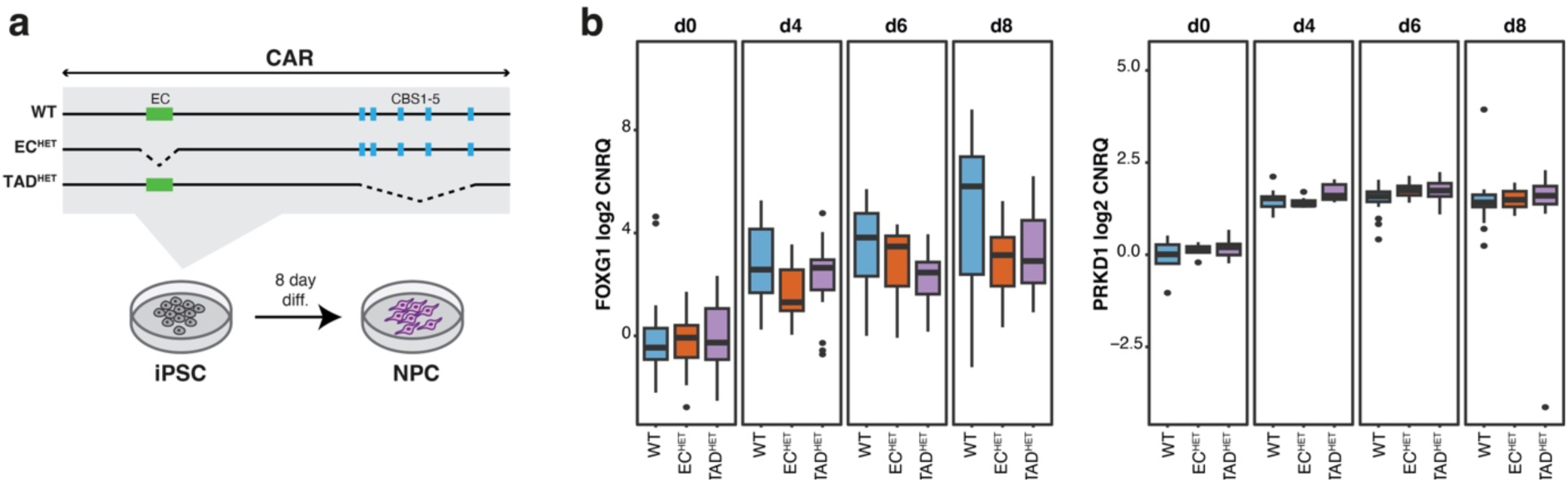
Deletion of the enhancer cluster (EC) or TAD boundary appears to decrease *FOXG1* expression during neural progenitor cell (NPC) differentiation. (a) Schematic overview of the generation of heterozygous knockout (KO) models for the EC and TAD boundary in iPSCs, through CRISPR using dual ribonucleoprotein (RNP) complexes, which are further differentiated towards NPCs. (b) RNA expression of *FOXG1* and *PRKD1* during an 8-day NPC differentiation for 3 genotypes: wildtype (WT), heterozygous deletion of the enhancer cluster (ECHET) and heterozygous deletion of the TAD boundary (TADHET). Each genotype encompasses at least 3 biological replicates (WT n=6; ECHET n=3; TADHET n=6). RNA expression data was normalized to reference gene expression (UBC, SDHA) and scaled to WT iPSCs.

Quantitative RT-qPCR analysis at day 0, 4, 6 and 8 of differentiation showed the induction of *FOXG1* and other NPC markers (PAX6) and the loss of pluripotency markers (*OCT3/4*) for all genotypes, confirming successful differentiation of all lines (**Fig 5b**, **Sup Fig S12**). Despite the induction of *FOXG1* RNA expression across all genotypes, there was a notable decrease in *FOXG1* expression at day 8 in heterozygous EC and TAD boundary deletion lines compared to wildtype controls (**Fig 5b**). Still, due to variability in *FOXG1* induction among wildtype lines, likely attributable to NPC population heterogeneity, this trend did not reach statistical significance. The data presented here therefore suggests that the presence of both the EC and TAD boundary within the CARR are crucial for maintaining normal *FOXG1* RNA expression levels. Yet to better quantify the effect, differentiation towards a more homogenous NPC population would be required.

## DISCUSSION

Through the introduction of novel sequencing technologies in the clinic and ensuing improved SV detection, there is an increasing number of cases in which putative causal SVs affecting non-coding sequences are reported^9^. However, interpreting the functional impact of these SVs remains challenging, given the incomplete annotation of the non-coding human genome and their potential impact on 3D chromatin structure. The 14q12 *FOXG1* region is a genomic locus recurrently affected by non-coding structural aberrations^16^. Through a comprehensive database and literature search, we assembled a cohort of 38 individuals with a *FOXG1* syndrome-like disorder, harbouring SVs that disrupt the *FOXG1* regulatory domain. No clear association was observed between phenotype severity and the type (deletion or BCA) or size of the structural variation. Notably, only one duplication was included in the cohort of *FOXG1* syndrome-like individuals. While two more duplications were present in the DECIPHER database overlapping the *FOXG1* TAD, we were not able to obtain any phenotypic information associated with these. Despite the diverse spectrum of SVs, with some CNVs affecting several megabases and encompassing multiple other protein-coding genes, *in silico* analysis using POSTRE^27^ identified *FOXG1* as the only candidate causative gene for the observed neurodevelopmental phenotype, reinforcing the hypothesis that aberrant *FOXG1* expression is the underlying mechanism responsible for the *FOXG1* syndrome-like disorder in this cohort. Several studies indeed reported aberrant FOXG1 expression in these cases^15,19,20,23^. Yet, expression levels were highly variable and assessed in sample types that may not adequately reflect FOXG1 regulation and expression in a neurodevelopmental context, such as blood, fibroblasts and, most often, LCLs. The reported expression differences (or lack thereof) should therefore be interpreted with care.

Through the identification of a novel ∼416kb *de novo* deletion in an individual with a clinical symptoms of *FOXG1* syndrome-like disorder, we could narrow down the previously identified smallest region of overlap to a 124 kb region downstream of *FOXG1,* that is disrupted in the entire *FOXG1*-like syndrome disorder cohort, here annotated as the CARR. We focused on characterizing the regulatory potential of the CARR, uncovering that it contains multiple cCREs, including a cluster of candidate enhancer elements and a TAD boundary consisting of five CBSs. Using UMI-4C in iPSCs, NSCs, NPCs and neurons we showed that the CARR strongly interacts with the *FOXG1* promoter during neurodevelopment and through *in vivo* enhancer assays in zebrafish embryos we validated neural enhancer activity for five cCREs within the EC. All identified enhancers (cCREa-e) drove tissue-specific GFP expression patterns in the zebrafish embryos, suggesting a role in regulating FOXG1 expression within specific developmental contexts^39–44^.

Subsequently, we opted to characterize the regulatory dynamics and impact of these elements within the CARR using NPCs. NPCs represent a highly suitable model for investigating *FOXG1* transcription and regulation, as FOXG1 is predominantly expressed in forebrain NPCs, where it orchestrates crucial processes like proliferation and differentiation^46^. The relevance of NPCs as a model extends to a direct link between NPC dysfunction and the phenotypic outcome of microcephaly observed in individuals with either *FOXG1* syndrome or *FOXG1* syndrome-like disorder, where reduced FOXG1 expression might lead to reduced expansion of the NPC population^46^. Epigenomic assays for active enhancer marks indicated that the EC region is dynamically activated during NPC differentiation, coinciding with the induction of *FOXG1* transcription. Finally, we observed a trend of decreased *FOXG1* transcription at day 8 of NPC differentiation upon heterozygous deletion of the EC and TAD boundary. Nevertheless this did not reach statistical significance due to variability in *FOXG1* induction levels associated with a heterogenous cell population obtained during NPC differentation. Although the widely used dual SMAD inhibition protocol is commonly assumed to generate a homogenous NPC population, our results as well as other studies suggest this is not always the case^39^. We observed expression of markers across anterior-posterior regions, including the forebrain (FOXG1, OTX2, SIX3 and DLX2), midbrain (LMX1A) and midbrain/hindbrain (EN1 and IRX3)^39^. Moreover, the *in vitro* derived NPCs used in our study represent early developmental stages of neural plate formation during neurulation, which occurs *in vivo* during the 3^rd^ and 4^th^ week of gestation^47^. This may account for the observed epigenomic differences between the *in vitro* NPCs and the *in vivo* human embryonic brain data. For example, while H3K27ac and H3K4me1 profiles from *in vitro* iPSC-derived NPCs confirmed the activation of the EC, they also revealed an additional candidate enhancer region immediately upstream of the EC, which had not been identified in the human embryonic brain DNase-seq data. Although we have yet to functionally characterize this newly discovered region, these findings emphasize the cell-type specific nature of regulatory elements like enhancers as previously shown in literature^48,49^. Follow-up analyses using more homogenous cell populations and allele-specific readouts will be required to better quantify the effect of regulatory deletions on *FOXG1* transcription. Overall, our results suggest a regulatory role for both the EC and TAD boundary within the CARR in establishing FOXG1 expression during neurodevelopment.

Despite the cell-type-specific activation of regulatory elements and expression of *FOXG1*, the TAD structure of the locus, including CTCF binding at the TAD boundary, appeared to be largely tissue-invariant. Such stable 3D chromatin structures have indeed been shown to be strongly associated with CTCF and cohesin binding and are thought to facilitate rapid enhancer-promoter communication since they ensure constant enhancer-promoter proximity, resulting in a more robust and efficient gene activation^50,51^. Using Hi-C and UMI-4C on case LCLs (del8), we have shown that deletion of the CARR and CBS cluster removes the conserved *FOXG1* TAD boundary and results in increased contacts with the distal TAD. Given the conserved TAD boundary structure and CTCF binding within this region, the effect on 3D chromatin structure would likely be the same in neuronal cells or tissues. Still, the mechanism of reduced *FOXG1* transcription upon deletion of the TAD boundary in NPCs remains unclear. Possibilities include a competition for interaction due to the increased contact of the *FOXG1* promoter with loci in the distal TAD, and/or the ectopic interaction with repressive elements.

Interestingly, in none of the individuals with a *FOXG1* syndrome-like disorder was the SV downstream of *FOXG1* restricted to the CARR. Even though identifying the smallest region of overlap has been used as a strategy to identify functional non-coding regions in the past^10,52–56^, it is important to consider that the *FOXG1*-like phenotype in these individuals likely arises from a cumulative effect caused by the deletion or translocation of multiple regulatory elements within the *FOXG1* TAD, rather than the removal of a single regulatory region. In line with this, we only observed a partial reduction of *FOXG1* transcription upon deletion of the EC. The observation that multiple regulatory elements are required to control a single gene, a phenomenon often referred to as redundancy, is not only plausible but also evolutionarily advantageous^57^ and protective against deleterious variants. Nevertheless, the smallest region of overlap approach remains valid to narrow down the search for functional non-coding elements and to enable more selective screening of the non-coding human genome.

In conclusion, this study delineates a commonly affected regulatory region (CARR) downstream of *FOXG1* that is disrupted in a cohort of individuals with *FOXG1* syndrome-like characteristics. By integrating epigenomic profiling, chromatin interaction analyses, genome editing, and functional assays, we identified and characterized an enhancer cluster and architectural elements within this region that play a role in regulating *FOXG1* transcription. These findings lay an important foundation for enhancing the interpretation of (non-coding) structural variants in individuals with *FOXG1* syndrome-like disorder.

## MATERIAL AND METHODS

### Cell culture

#### Lymphoblastoid cell lines (LCLs)

LCLs were cultured in RPMI (Roswell Park Memorial Institute). RNA was extracted using the Maxwell® RSC instrument (AS4500, Promega) using the Maxwell® RSC simply RNA Tissue kit (AS1340, Promega).

#### Induced pluripotent stem cells (iPSCs)

Human iPSC cells (Axol Bioscience, lot #30270222072019) were cultured in feeder-free conditions on Geltrex® LDEV-Free hESC-Qualified Reduced Growth Factor Basement Membrane Matrix (Fisher Scientific #12063569)-coated 6-well plates in mTeSR Plus medium (Stemcell Technologies #100-0276) that was changed every other day. Cells were dissociated with ReLeSR (Stemcell Technologies #100-0483) after reaching 80% confluency.

#### Neural stem cells (NSCs) & Induced neurons (iNs)

To minimize technical artifacts, hiPSCs were plated as single cells at 80% confluence on a Matrigel-coated 6-well plate with Y-27632 dihydrochloride (1 mg/mL). Polybrene was added at 8 mg/mL 1 h after re-plating. Cells were incubated with polybrene for 10–15 min prior to the addition of lentivirus. Lentiviral constructs for directed differentiation of hiPSCs into iNs were made as described previously^58^. hiPSC lines were differentiated into NSCs with the protocol detailed by Thermo Fisher (MAN0008031)^59^.

#### Neural progenitor cells (NPCs)

iPSCs were differentiated towards NPCs using the STEMdiff™ SMADi Neural Induction Kit following the monolayer protocol provided by the manufacturer (Catalog #08581), which directs differentiation by blocking TGF-β/BMP-dependent SMAD signaling. NPCs were seeded at a density of 4 × 10^5^ cells per well in a Geltrex coated 12-well plate in STEMdiff™ SMADi Neural Induction medium over 8 or 12 days, with media changes each day. For the 12 day protocol the cells were split at day 6, for the 8 day protocol no split was performed.

### DNA extraction

Total DNA was extracted from iPSCs and iPSC-derived NPCs at day 8 of differentiation. The cells were rinsed twice with D-PBS and harvested with a cell scraper. The PBS-containing cells were centrifuged (200 x g for 5 min), the supernatant was discarded and the pellet was resuspended in 200μL D-PBS. DNA was obtained from this solution using the QiaAmp DNA Blood Mini kit (Qiagen #51106) on the Maxwell rapid Sample Concentrator (RSC) which allows automated extraction of DNA from different type of cells.

### CNV-seq and breakpoint finemapping

Molecular karyotyping was performed through low-pass whole genome sequencing on genomic DNA (gDNA). Whole genome libraries were prepared with the Illumina DNA PCR-Free Prep kit (20041795, Illumina) according to the manufacturer’s instructions. The libraries were subsequently paired-end sequenced (2×50 bp) on a NovaSeq6000 system (Illumina) to a depth of 75M reads. The raw sequencing data was then aligned to the human reference genome GRCh38/hg38 with bowtie2^60^. Further processing and analyses were done using WisecondorX^61^ and Vivar^62^. Breakpoints were then finemapped at base pair resolution through CNV-qPCR^63^, followed by PCR amplification over the putative breakpoints and Sanger sequencing of the amplicon.

### RNA isolation, cDNA synthesis and quantitative PCR

Total RNA was extracted using QIAzol Lysis reagent (Qiagen #79306) and the Direct-zol RNA Miniprep Kit (Zymo Research #R2052). Complementary DNA (cDNA) synthesis was performed using the iScript cDNA synthesis kit (Bio-Rad #170-8891; Bio-Rad, Hercules, California, USA). Quantitative polymerase chain reaction (qPCR) was performed using 2× SYBR Green SsoAdvanced Supermix (Bio-Rad #172-5274) with 5 μM forward and reverse primer and 5 ng cDNA input. Expression values were analyzed using qBase + software (Biogazelle, Ghent, Belgium) and normalized using two reference genes (*UBC, SDHA*).

### Generation of in vivo reporter constructs and functional characterization of cCREs in zebrafish

The cCRE sequences were synthesized as gBlocks (IDT) with 30 bp flanking regions designed to overlap with the ends generated by AscI and PacI restriction digestion of the E1b vector^37^. The digested E1b vector and gBlocks were assembled using Gibson assembly (Gibson Assembly Master Mix, NEB # E2611L). In parallel, the same gBlocks were cloned into a TOPO vector via A-tailing. The TOPO vector, containing Gateway recombination sites, was then used for Gateway cloning into the ZED vector^38^ using the Gateway LR Clonase II enzyme mix (Invitrogen #11791-020). E1b contains a minimal promoter followed by a GFP reporter gene. ZED, in addition, includes a transgenesis internal control cassette comprising the cardiac actin promoter driving the expression of red fluorescent protein (RFP) and insulator sequences to mitigate position effect artifacts. The combination of these vectors enables a more accurate assessment of enhancer functionality, as each provides complementary insights into enhancer-driven gene expression. These vectors were microinjected into one-cell stage zebrafish embryos along with Tol2 transposase mRNA for genomic integration. As a readout, GFP fluorescence was observed and its localization was annotated at 1, 2, and 3 days post fertilization (dpf) to evaluate enhancer activity.

### Identification of candidate cis-regulatory elements (cCREs)

A set of cCREs with enhancer-like epigenomic properties was created by overlapping entries in the Annotated Regulatory Index (DHS) by Meuleman et al. (meuleman.org)^33^ and enhancer-annotated entries in the ENCODE SCREEN v3 dataset^34^ using the GenomicRanges R package. To obtain a complete list of cCREs with accessibility in neuronal tissues, we selected peaks defined by Meuleman et al. as “tissue irrelevant” or “nervous”, showing accessibility in 10% of relevant samples (Organ = “Neural”/”Brain”/”Germ”). Finally, enhancer-like cCREs located within the same TAD as *FOXG1* were selected, based on TAD boundaries determined using brain Hi-C data by Rahman et al.^32^

### UMI-4C library preparation and analysis

UMI-4C was performed according to the protocol by Schwartzman et al.^30^ In brief, 1 (iNs), 5 (iPSCs, NSCs, NPCs) or 10 (LCLs) million cells per sample were detached, resuspended in a PBS/10% FBS solution and crosslinked using formaldehyde (final concentration 2%) (Sigma-Aldrich, F1635-500ML) for 10 min. The reaction was quenched on ice using glycine (final concentration 0.125M). Cells were then centrifuged, washed and resuspended in cold lysis buffer (50mM Tris-HCl, 150mM NaCl, 5mM EDTA, 0.5% NP-40, 1% Triton X-100 and 1 tablet protease inhibitor per 10mL (cOmplete, Mini, Sigma-Aldrich, 11836153001)) and incubated for 10 min. After centrifugation, pellets were washed, snap-frozen in liquid nitrogen and stored at −80°C until further processing. Frozen pellets were resuspended in 500µL pre-diluted DpnII buffer and 15µL pre-heated 10% SDS and incubated on a thermomixer for 1h at 37°C, shaking at 900 RPM. We halved these volumes and those mentioned below for smaller cell pellets (from 1 million cells). After adding 150µL 10% Triton X-100, the solution was incubated again (1h, 37°C, 900 RPM). We digested the chromatin using 600U DpnII (NEB, R0543L) in three stages (200U for 2h, 200U overnight, 200U for 2h) at 37°C and 900 RPM. The solution was incubated at 65°C for 20 min to inactivate the restriction enzyme and put on ice. Next, the chromatin was ligated by adding 4000U of T4 DNA ligase (NEB, M0202M) and 10x T4 DNA ligase buffer to a total volume of 1300µL and incubating the solution overnight at 16°C and 300 RPM. Ligated chromatin was de-crosslinked by incubation with 8µL proteinase K (20mg/mL, Qiagen, 19131) (overnight, 65°C, 300 RPM). A 3C template was then purified using 1x Ampure XP beads (Beckman Coulter, A63881).

Up to 4µg 3C template per sample was sheared using microTUBE snap-cap tubes (Covaris, 520045) in a Covaris M220 sonicator to an average fragment length of 300bp. UMI-4C sequencing libraries were generated using the NEBNext Ultra II library prep kit (NEB, E7645L). For each sample, library prep was performed in four parallel reactions with a maximum input of 1000ng per reaction, including an AmpureXP beads size selection targeting fragments with a length of 300-400bp (unless input was <100ng) and 4-8 cycles of PCR enrichment depending on the input. Next, two nested PCR reactions were performed to enrich for fragments captured by the viewpoint of interest, both using 2µL 10mM Illumina enrichment primer 2 and either a viewpoint-specific “upstream” (reaction 1, 2µL 10mM) or “downstream” (reaction 2, 2µL 10mM) primer (**Sup Table S4**). For each sample, we performed up to 8 nested PCR reactions in parallel with an input of 100-200ng per reaction using the KAPA2G Robust ready mix (Sigma-Aldrich, KK5702) in a volume of 50µL. PCR program: 3 min 95°C, 20 cycles (18 cycles for reaction 2) of 15 sec 95°C, 15 sec 55°C, 60 sec 72°C and final elongation of 5 min 72°C. Between PCR reactions the product was cleaned up using 1x AmpureXP beads and eluted in 21µL. The final PCR product was cleaned up using 0.7x AmpureXP beads and eluted in 25µL. Reactions per sample were pooled and library concentration was quantified via qPCR using the KAPA SYBR FAST qPCR Master Mix (Sigma-Aldrich, KK4602).

UMI-4C libraries were pooled and sequenced on an Illumina HiSeq or NovaSeq (paired-end, 2×150 cycles). Reads were first filtered based on the presence of the downstream primer sequence (20% mismatch allowed). The resulting fastq files were used as input to the UMI-4C R package (https://github.com/tanaylab/umi4cpackage) to generate genomic interaction tracks representing UMI counts (i.e. unique interactions) per genomic restriction fragment. The package was then used to generate smoothed, viewpoint-specific interaction profiles for the region of interest. For each profile, interaction counts were normalized to the total UMI count within the profile.

### Hi-C library preparation and analysis

Case LCLs were crosslinked, lysed and snapfrozen according to the protocol mentioned above under ‘UMI-4C’. Crosslinked nuclei were then used to construct Hi-C libraries, following the in situ Hi-C protocol adopted by the 4D Nucleome consortium^31^ with a few adaptations, as previously described by D’haene et al.^64^ Briefly, for each replicate ∼5 million pre-lysed, crosslinked nuclei were digested overnight using 250 U DpnII restriction enzyme (New England Biolabs, R0543L). DNA ends were marked by incorporating biotin-14-dATP (Life Technologies, 19524-016) and ligated for 4 hours using 2000 U T4 DNA ligase (New England Biolabs, M0202L). Subsequently, crosslinks were reversed overnight using proteinase K (Qiagen, 19131) and Hi-C template DNA was purified using 1x AMPure XP beads (Beckman Coulter, A63881) and stored at 4°C until library preparation. Hi-C template DNA was sheared to a size of 300-500 bp using microTUBE snap-caps (Covaris, 520045) in a Covaris M220 sonicator and MyOne Streptavidin T1 beads (Life Technologies, 65601) were used to pull down biotinylated ligation junctions. Next, samples were split into 5 µg aliquots for sequencing library preparation using the NEBNext Ultra II DNA Library Prep Kit (New England Biolabs, E7645L) and NEBNext Multiplex Oligos (New England Biolabs, E7335L). Amplified libraries were purified and size selected using 0.55x and 1.2x AMPure XP beads (Beckman Coulter, A63881). Pooled libraries were sequenced on an Illumina NovaSeq 6000 using 100 bp paired-end reads to a depth of ∼500 million reads per sample.

FASTQ files containing raw sequencing data were processed into Hi-C contact matrices containing both raw and Knight-Ruiz (KR) normalized counts using the Juicer pipeline (v1.6)^65^ with BWA-MEM mapping (v0.7.17)^66^ to the hg38 reference genome. A processed and normalized Hi-C matrix for control LCLs was obtained from Melo *et al.*^13^ Insulating boundaries between self-interacting domains were identified based on diamond insulation score minima. We used cooltools^67^ to calculate a genome-wide contact insulation score with 250 kb window size for KR normalized Hi-C contact matrices (MAPQ>30) at 25 kb resolution. Insulating boundaries were determined by applying automated ‘Li’ thresholding (from the scikit-image Python package) on boundary strength. We used FAN-C^68^ to visualize KR normalized case vs. control Hi-C matrices and fold-change matrices for the region of interest.

### Generating CRISPR Cas9 engineered iPSC lines

#### Design of gRNA’s

The gRNAs were designed making use of the Custom Alt-R™ CRISPR-Cas9 guide RNA IDT design tool. For the KO of both the EC and TAD, two gRNAs (IDT) were designed within the region 1kb region upstream and downstream of the target region. The gRNAs with the highest on-target potential and lowest off-target risk were selected (**Sup Table S5**).

#### Formation of RNP complex

The crRNA and tracrRNA (IDT) were resuspended in IDTE buffer to final concentrations of 200 µM each. These two RNA oligonucleotides were mixed in equimolar concentrations to a final duplex concentration of 100 µM. This mixture was heated at 95°C for 5 min and afterwards cooled to room temperature for approximately 15 minutes and then placed on ice. For each well undergoing electroporation, the crRNA:tracrRNA duplex (1,01µl per reaction) and Alt-R Cas9 enzyme (IDT) components (61µM stock; 1,43µl per reaction) were diluted in PBS (1,77µl per reaction). This mixture was incubated at room temperature for 20 minutes and stored at 4°C after incubation.

#### Electroporation of RNP complex into iPSCs

Electroporation was performed using the Lonza® 4D-Nucleofector™ X Unit with a P3 primary cell 4D Nucleofector X kit.

Cells were collected during their exponential growth phase. The remaining medium was aspirated from the well and 1 mL of Accutase (STEMCELL Technologies) was added to each well of a 6-well plate. The plate was incubated at 37°C and 5% CO2 for approximately 5 minutes, or until colonies appeared to be dissociated. Cells were gently washed from the surface of the plate with 2ml mTeSR Plus (STEMCELL Technologies) using a pipettor fitted with a 1000 µl tip, pipetting the suspension up and down 2-3 times to break up small aggregates into single cells necessary for the electroporation. The cell suspension was transferred to a 15 mL conical tube and centrifuged at 300 x g for 5 minutes. The supernatant was aspirated, being careful not to disturb the cell pellet. Cells were resuspended in at least 2 mL of Single-Cell Plating Medium (mTeSR Plus and Clone R ((STEMCELL Technologies) in a 1:10 dilution, used to improve single-cell survival) and mixed by flicking the tube 2-3 times.

Cells were counted using an automated cell counter (Cellometer Auto T4). For each electroporation condition (f.e. EC, TAD, GFP, no nucleofection) 3 x 10^5 cells were added to a new 15 mL conical tube. The supernatant was removed without disturbing the pellet, and the cells were washed in 1 mL 1x PBS. Cells were centrifuged at 400 x g for 10 minutes and the supernatant was removed. Cells (300,000 cells/reaction) were resuspended by adding Nucleofector Solution and supplement to the Nucleofector solution (4.5:1 ratio – solution)). For each condition, 17 µl of the prepared solution was pipetted into a sterile microcentrifuge tube. For each condition (EC, TAD and GFP), 4.21 µl of each crRNA:tracrRNA (for the EC and TAD both the upstream and downstream complexes are added, total 8.42 µl) and 0.84 µl (4 µM) of 100 µM Alt-R Cas9 Electroporation Enhancer (IDT) (total volume = ±26 µl) were added. The mixture was pipetted up and down and 22 µl of the mixture was transferred to the wells of the Nucleocuvette. The Nucleocuvette was gently tapped to ensure no air bubbles were present. The Nucleocuvette was placed in the device and the electroporation program CA-137 (P3 solution) was started. After electroporation, the Nucleocuvette was removed from the instrument, and 80 µl of pre-warmed culture media was added per well. Cells were resuspended by gently pipetting up and down, and transferred to a Geltrex coated 12-well plate, containing Single-Cell plating medium. A full medium change was performed after 24h with mTeSR Plus. These edited iPSC were further expanded and kept in culture following standard iPSC culturing protocols.

#### Screening edited iPSCs in bulk

Successful editing was assessed in the bulk of the cells via DNA isolation and amplification of the target region, followed by targeted next-generation-sequencing (NGS). For larger deletions, both primers outside and inside the deletion were designed (**Sup Fig S13**, **Sup Table S6**). If successfully edited clones were present within the bulk, a PCR fragment was expected in all three PCR reactions. These were further kept in culture and expanded to perform serial limiting dilution to obtain monoclonal edited iPSC clones. Similar PCR reactions as described above were performed on the extracted DNA of these clones.

#### Limiting dilution

To obtain clonal cell lines, the transfected iPSCs were single cell isolated through serial limiting dilutions. Once the bulk cells reached their exponential growth phase, they were dissociated into single cells using Accutase (STEMCELL Technologies). To prevent clumping of cells, a cell strainer was used. The cells were diluted to a final concentration of 0.5 cells per 100 µl of Single-Cell plating medium, to reduce the likelihood of having multiple cells per well. 100 µl of diluted cells were added to each well of a 96-well plate, coated with Geltrex. After four to eight days, individual colonies were observed and further used for expansion and screening.

#### Screening edited iPSC clones

To examine the successful deletion and assess monoclonality of the cloned hiPSCs, genome DNA was extracted using QuickExtract DNA isolation kit (QE09050, Epicentre). Cells were dissociated using ReLeSR (STEMCELL technologies) and resuspended into 100 µl of mTeSR Plus + 10 µM Rock Inhibitor. Half of the cells were transferred (∼50 µL) to 96 well plate containing mTesR Plus + 10 µM Rock Inhibitor for further cell expansion. The other half (∼50 µL) was transferred to a 96 well PCR plate, while maintaining the location of each sample, for genomic DNA extraction. The PCR plate was spun down at 3800 rpm for 30 minutes at 4°C, supernatants was removed and 30 µl of the QuickExtract DNA solution was added to the pellet and protocol was performed further according to the manufacturer’s instructions. Similar PCR reactions as described above were performed on the extracted DNA of these clones. Finally, monoclonality was confirmed via DNA isolation and amplification of the target region, followed by targeted NGS (indels) and CNV-seq (kb-sized deletions) (**Sup Fig S14**).

### CUT&RUN library preparation

On day 0, 4, 6 and 8 of differentiation, iPSCs/NPCs were washed twice with room temperature 1X PBS. Cells were lifted from the dish using a cell lifter and gently resuspended using a P1000 pipet. Cells were counted using an automated cell counter, and the appropriate number of cells was taken. For each condition 5 x 10^5 cells were required, however it is recommended to batch process and therefore cells for all conditions were pooled into a 15ml falcon tube. The lysate was transferred to a fresh 15ml falcon tube and spun down at 600 g at RT for 3 min. The supernatant was carefully discarded and nuclei were isolated by addition of Nuclei Extraction Buffer (NEB, 20 mM Hepes (KOH) pH 7.9, 10 mM KCl, 0.5 mM Spermidine, 0.1% Triton X-100, 20% Glycerol, 1mM MnCl2, 1 tablet cOmplete MINI EDTA-free Protease inhibitor (Roche)) (100µl per condition). The samples were incubated for 10 minutes on ice and spun down at 600 x g at 4°C for 3 minutes. Supernatants was removed and 100µl NEB was added per condition.

For these first steps it is recommended to work in batch, pool everything together for all samples of the same datapoint or cell type. Per reaction, 10 μl/sample Concavalin-A conjugated beads were activated by washing twice in 100 μl/sample cold Bead Activation Buffer (20 mM HEPES-KOH, pH 7.9, 10 mM KCl, 1 mM CaCl2, 1 mM MnCl_2_) on a magnetic stand and then resuspended in 11 μl/sample Bead activation Buffer. Freshly 100 μl/sample extracted nuclei were added to the activated beads and incubated at 4°C for 10 min with rotation. Beads-bound nuclei were placed on a magnetic stand and resuspended in Antibody buffer 50 μl/sample (2mM EDTA, 1 tablet cOmplete MINI EDTA-free Protease inhibitor (Roche) in wash buffer). Primary antibodies were added to the beads-bound nuclei and gently mixed, see **table S7** (final dilution of 1:50 or 1:100 in 50 μl volume). Mixtures were incubated with rotation at 4°C overnight on a nutator.

After washing twice on a magnetic stand in 250 μl/sample Washing Buffer (WB, 20 mM Hepes pH 7.5, 150 mM NaCl, 0.5 mM Spermidine, 1 tablet cOmplete EDTA-free Protease inhibitor), nuclei were resuspended in 50 μl/sample cold Washing Buffer. 2.5 μl of pAG-MNAse (15-1016-EPC Epicypher) was added to the mix and incubated with rotation for 10 min at 4 °C. Next, beads were washed twice in 250 μl/sample cold Washing Buffer on a magnetic strand and resuspended in 50 μl cold Washing Buffer. The samples were equilibrated to 0°C in an ice bath and 1 μl/sample of 100 mM CaCl2 were added to the reaction. After an incubation of 2h at 4°C on a nutator, 33 μl/sample 2X STOP buffer (340 mM NaCl, 20 mM EDTA, 4 mM EGTA, 50 μg/ml RNAseA, 50 μg/ml Glycogen, 10 ng/ml spike-in DNA) was added and beads were incubated for 10 min at 37°C. The supernatants was extracted and DNA was purified using the CUTANA™ Quick Cleanup DNA Purification Kit, following manufacturer’s instructions. DNA concentration was measured with Qubit (Thermo Fisher Scientific).

Libraries were prepared using the NEBNext Ultra II Library prep Kit (E7645S, New England Biolabs) according to the manufacturer’s protocol using half of the volumes. Adapters were used at a working concentration of 15 μM, 1.5 μM, 0.6 μM based on the DNA input. A cleanup of the adaptor-ligated DNA without size selection was performed making use of AMPure XP beads (1.8X). Libraries (13 μL) were PCR amplified with unique indexes (15 μL NEBNext Ultra II Q5 Master Mix and 1 μL i7 Primer) (New England Biolabs) for 8-15 cycles depending on the start DNA input.

A cleanup of the PCR reaction with size selection was performed making use of AMPure XP beads for the selection of fragments 150bp(0.5X) < DNA fragments < 500bp(1.2X). After cleanup, fragment size and concentration was analyzed by Fragment Analyzer. CUT&RUN library concentrations were measured with the Illumina Kapa Library quantification kit (Roche #07960140001). All pre-made libraries were sequenced at on Illumina NEXTseq2000 platform (150 bp paired-end sequencing). Each replicate was sequenced at a sequencing depth of 10 million reads.

### CUT&RUN analysis

Read processing and peak calling was performed using a dedicated nextflow CUT&RUN pipeline available through nf-core^69^ (https://doi.org/10.5281/zenodo.5653535). Briefly, raw reads were trimmed, QC filtered and aligned to the hg38 reference genome using Bowtie2 (v2.4.4)^60^. Upon Q-score filtering, duplicate removal and CPM normalization, peaks in target samples were called with SEACR (v1.3)^70^ using corresponding IgG samples as controls. A consensus peak list per sample was obtained by requiring the presence of peaks in both replicates.

### Reanalysis of single-cell RNA-seq data for cell-type-specific expression analysis of *FOXG1*

Single-cell RNA-seq data of the developing human brain (Braun et al.^71^) was accessed through GitHub (https://github.com/linnarsson-lab/developing-human-brain) in h5ad format with CELLxGENE annotations (<u>human_dev.h5ad</u>). The single-cell RNA-seq data from the human neural organoid cell atlas HNOCA^45^ was sourced from CELLxGENE (https://cellxgene.cziscience.com/collections/de379e5f-52d0-498c-9801-0f850823c847). The datasets were subset and heatmaps were generated in python using the heatmap function from the seaborn package.

## DATA AVAILABILITY

All datasets generated in this study will be available through the Gene Expression Omnibus (GEO) repository upon publication.

## COMPETING INTERESTS

The authors declare no competing interests.

## ETHICS

This study was conducted in accordance with ethical guidelines and received approval from the Ethics Committee (EC) under approval number ECD 20-86, ONZ-2022-0100 and ECD 2019/1430. Informed consent was obtained from the patient prior to data collection, ensuring they fully understood the purpose of the study and the potential for their data to be made publicly available. All data were anonymized to protect patient confidentiality and comply with relevant data protection regulations.

## AUTHORS’ CONTRIBUTIONS

E.D., B.M. and S.V. conceived the overall project. E.D., L.H. and S.V. designed the experiments. B.C. examined the patient, provided biological samples and gathered clinical information. H.G. contributed additional clinical information. A.D. and B.M. provided cytogenetic analysis and molecular diagnosis. E.D., L.H., N.V.L., M.B.V., M.d.R.P.B., E.D. and L.V. performed functional experimental assays. E.D., S.L. and L.C. performed bioinformatic analyses. E.D., L.H. and S.V. interpreted the results and wrote the manuscript. All authors reviewed and approved the final version of the manuscript.

## FUNDING

Financial support has been provided by grants 1520518N and G055422N from the Research Foundation – Flanders (FWO) and BOF/STA/201909/009 from the Special Research Fund (BOF) from Ghent University. E.D. is supported by a postdoctoral grant from the Research Foundation Flanders (FWO 12D8523N) and was previously supported by Marguerite-Marie Delacroix and FWO doctoral fellowships. L.H. and N.V.L. are supported by a doctoral fellowship from the Research Foundation Flanders (L.H.:FWO 11I2525N & N.V.L.: FWO 11A4C25N) and were previously supported by a Marguerite-Marie Delacroix doctoral fellowship. M.d.R.P.B. was and L.C. is supported by a Marguerite-Marie Delacroix doctoral fellowship.

## Data Availability

All in vitro data produced in this work will be made available in public repositories. The personal data is stored on the internal servers of the Center for medical genetics but due to restrictions, the personal data cannot be shared.

## ACKNOWLEDGEMENTS

We extend our gratitude to the participating individuals and their families for their contribution in this study. Crosslinked cell pellets from human NSCs and neurons were generously provided by dr. K. Mohajeri-Stickels and prof. M. Talkowski. Hi-C interaction matrices from control LCLs were kindly provided by dr. U. Melo and prof. S. Mundlos. We thank the Core Zebrafish Facility Ghent (ZFG) and dr. Andy Willaert for their expert technical assistance with *in vivo* enhancer assays. The E1b vector and the ZED vector were generously provided by prof. R.Y. Birnbaum and dr. J. Bessa, respectively.

This study makes use of data generated by the DECIPHER community. A full list of centres who contributed to the generation of the data is available from https://deciphergenomics.org/about/stats and via email from. DECIPHER is hosted by EMBL-EBI and funding for the DECIPHER project was provided by the Wellcome Trust [grant number WT223718/Z/21/Z]. We would specifically like to thank prof. Alessandra Renieri, dr. Margherita Baldassarri, dr. Chiara Fallerini, dr. Edward Blair, dr. Deborah Shears and dr. Pernille M. Torring for their kind cooperation and providing information on cases included in the DECIPHER database.

## SUPPLEMENTARY MATERIAL

### Supplementary Text. Clinical description of new patient included in the manuscript (not included on MedRxiv)

**Supplementary Figure S1. Identification and finemapping of patient deletion.**

**Supplementary Figure S2. Overview of all CNVs observed in patients overlapping the bigger FOXG1 TAD based on the DECIPHER database.**

Supplementary Figure S3. Detailed view of the enhancer cluster and TAD boundary. Supplementary Figure S4. Chromatin interaction frequencies at the *FOXG1* and *PRKD1* locus. Supplementary Figure S5. TAD structure conservation at the FOXG1 locus across tissues.

**Supplementary Figure S6. CTCF binding at the commonly affected regulatory region (CARR) across ENCODE tissues and cell types.**

**Supplementary Figure S7. RAD21 binding at the commonly affected regulatory region (CARR) across ENCODE tissues and cell types.**

**Supplementary Figure S8. Quantification of zebrafish enhancer assay results.**

**Supplementary Figure S9. Single-cell RNA-seq data from human brain development shows high FOXG1 expression in ventral telencephalic NPC and neurons.**

**Supplementary Figure S10. Characterization of NPC populations using regional markers. Supplementary Figure S11. Regulatory dynamics of NPC markers during NPC differentiation. Supplementary Figure S12. Successful differentiation of WT and CRISPR KO models for the EC and TAD boundary towards NPCs.**

**Supplementary Figure S13. Schematic overview of PCR reactions to assess successful generation of the heterozygous KO models.**

**Supplementary Figure S14. Assessing monoclonality and successful editing of the clones within the different genotypes.**

**Supplementary Table S1. Overview of FOXG1 syndrome-like cases harbouring non-coding sturctural variants (SVs) overlapping the FOXG1 topologically associating domain (not affecting FOXG1).**

**Supplementary Table S2. Results of POSTRE in silico analysis of structural variants (SVs).**

**Supplementary Table S3. Results of zebrafish enhancer assays for candidate cis-regulatory elements (cCREs).**

**Supplementary Table S4. Primer design for UMI-4C.**

**Supplementary Table S5. Paired CRISPR/Cas9 sgRNA design for engineering enhancer cluster (EC) and TAD boundary (TAD) deletions.**

**Supplementary Table S6. Primer design for PCR screening of CRISPR/Cas9 edited clones. Supplementary Table S7. Antibodies used for CUT&RUN assays.**

## Notes

### Competing Interest Statement

The authors have declared no competing interest.

### Author Declarations

Ethics committee of Ghent University Hospital gave ethical approval for this work.

## REFERENCES

1. Hou, P.-S., hAilín, D. Ó., Vogel, T. & Hanashima, C. Transcription and Beyond: Delineating FOXG1 Function in Cortical Development and Disorders. Front. Cell. Neurosci. 14, (2020).

2. Chen, D. et al. Loss of Foxg1 Impairs the Development of Cortical SST-Interneurons Leading to Abnormal Emotional and Social Behaviors. Cereb. Cortex N. Y. N 1991 29, 3666–3682 (2019).

3. Zhu, W. et al. Precisely controlling endogenous protein dosage in hPSCs and derivatives to model FOXG1 syndrome. Nat. Commun. 10, (2019).

4. Mariani, J. et al. FOXG1-Dependent Dysregulation of GABA/Glutamate Neuron Differentiation in Autism Spectrum Disorders. Cell 162, 375–390 (2015).

5. Won, H. et al. Chromosome conformation elucidates regulatory relationships in developing human brain. Nature 538, 523–527 (2016).

6. Mitter, D., et al. FOXG1 Syndrome: GenotypePhenotype Association in 84 Patients with FOXG1 Variants. Neuropediatrics 48, S1–S45 (2017).

7. Gold, W. A., Krishnarajy, R., Ellaway, C. & Christodoulou, J. Rett Syndrome: A Genetic Update and Clinical Review Focusing on Comorbidities. ACS Chem. Neurosci. 9, 167–176 (2018).

8. Kortum, F. et al. The core FOXG1 syndrome phenotype consists of postnatal microcephaly, severe mental retardation, absent language, dyskinesia, and corpus callosum hypogenesis. J. Med. Genet. 48, 396–406 (2011).

9. D’haene, E. & Vergult, S. Interpreting the impact of noncoding structural variation in neurodevelopmental disorders. Genet. Med. Off. J. Am. Coll. Med. Genet. 23, 34–46 (2021).

10. Mehrjouy, M. M. et al. Regulatory variants of FOXG1 in the context of its topological domain organisation. Eur. J. Hum. Genet. EJHG 26, 186–196 (2018).

11. Poszewiecka, B. et al. TADeus2: a web server facilitating the clinical diagnosis by pathogenicity assessment of structural variations disarranging 3D chromatin structure. Nucleic Acids Res. 50, W744–W752 (2022).

12. Lu, G. et al. Identification of a de novo mutation of the FOXG1 gene and comprehensive analysis for molecular factors in Chinese FOXG1-related encephalopathies. Front. Mol. Neurosci. 15, (2022).

13. Melo, U. S. et al. Hi-C Identifies Complex Genomic Rearrangements and TAD-Shuffling in Developmental Diseases. Am. J. Hum. Genet. 106, 872–884 (2020).

14. Pienkowski, V. M. et al. Mapping of breakpoints in balanced chromosomal translocations by shallow whole-genome sequencing points to EFNA5, BAHD1 and PPP2R5E as novel candidates for genes causing human Mendelian disorders. J. Med. Genet. 56, 104–112 (2019).

15. Allou, L. et al. 14q12 and severe Rett-like phenotypes: new clinical insights and physical mapping of FOXG1-regulatory elements. Eur. J. Hum. Genet. 20, 1216–1223 (2012).

16. Redin, C. et al. The genomic landscape of balanced cytogenetic abnormalities associated with human congenital anomalies. Nat. Genet. 2016 491 49, 36–45 (2016).

17. Craig, C. P. et al. Diagnosis of FOXG1 syndrome caused by recurrent balanced chromosomal rearrangements: case study and literature review. Mol. Cytogenet. 13, 40 (2020).

18. Ibn-Salem, J. et al. Deletions of chromosomal regulatory boundaries are associated with congenital disease. Genome Biol. 15, 423 (2014).

19. Ellaway, C. J. et al. 14q12 microdeletions excluding FOXG1 give rise to a congenital variant Rett syndrome-like phenotype. Eur. J. Hum. Genet. EJHG 21, 522–527 (2013).

20. Takagi, M. et al. A 2.0 Mb microdeletion in proximal chromosome 14q12, involving regulatory elements of FOXG1, with the coding region of FOXG1 being unaffected, results in severe developmental delay, microcephaly, and hypoplasia of the corpus callosum. Eur. J. Med. Genet. 56, 526–528 (2013).

21. Shoichet, S. A. et al. Haploinsufficiency of novel FOXG1B variants in a patient with severe mental retardation, brain malformations and microcephaly. Hum. Genet. 117, 536–544 (2005).

22. Alosi, D. et al. Dysregulation of FOXG1 by ring chromosome 14. Mol. Cytogenet. 8, 24 (2015).

23. Goubau, C. et al. Platelet defects in congenital variant of Rett syndrome patients with FOXG1 mutations or reduced expression due to a position effect at 14q12. Eur. J. Hum. Genet. EJHG 21, 1349–1355 (2013).

24. Firth, H. V. et al. DECIPHER: Database of Chromosomal Imbalance and Phenotype in Humans Using Ensembl Resources. Am. J. Hum. Genet. 84, 524–533 (2009).

25. Visel, A., Minovitsky, S., Dubchak, I. & Pennacchio, L. A. VISTA Enhancer Browser--a database of tissue-specific human enhancers. Nucleic Acids Res. 35, D88–92 (2007).

26. Spielmann, M., Lupiáñez, D. G. & Mundlos, S. Structural variation in the 3D genome. Nat. Rev. Genet. 19, 453–467 (2018).

27. Sánchez-Gaya, V. & Rada-Iglesias, A. POSTRE: a tool to predict the pathological effects of human structural variants. Nucleic Acids Res. 51, e54 (2023).

28. Sifrim, A. et al. Distinct genetic architectures for syndromic and nonsyndromic congenital heart defects identified by exome sequencing. Nat. Genet. 48, 1060–1065 (2016).

29. Alter, S. et al. Telangiectasia-ectodermal dysplasia-brachydactyly-cardiac anomaly syndrome is caused by de novo mutations in protein kinase D1. J. Med. Genet. 58, 415–421 (2021).

30. Schwartzman, O. et al. UMI-4C for quantitative and targeted chromosomal contact profiling. Nat. Methods 13, 685–691 (2016).

31. Rao, S. S. P. et al. A 3D map of the human genome at kilobase resolution reveals principles of chromatin looping. Cell 159, 1665–1680 (2014).

32. Rahman, S. et al. Lineage specific 3D genome structure in the adult human brain and neurodevelopmental changes in the chromatin interactome. Nucleic Acids Res. 51, 11142–11161 (2023).

33. Meuleman, W. et al. Index and biological spectrum of human DNase I hypersensitive sites. Nature 584, 244–251 (2020).

34. Moore, J. E. et al. Expanded encyclopaedias of DNA elements in the human and mouse genomes. Nature 583, 699–710 (2020).

35. Dimitrieva, S. & Bucher, P. UCNEbase—a database of ultraconserved non-coding elements and genomic regulatory blocks. Nucleic Acids Res. 41, D101–D109 (2013).

36. Hébert, J. M. & McConnell, S. K. Targeting of cre to the Foxg1 (BF-1) locus mediates loxP recombination in the telencephalon and other developing head structures. Dev. Biol. 222, 296–306 (2000).

37. Birnbaum, R. Y. et al. Coding exons function as tissue-specific enhancers of nearby genes. Genome Res. 22, 1059–1068 (2012).

38. Bessa, J. et al. Zebrafish enhancer detection (ZED) vector: A new tool to facilitate transgenesis and the functional analysis of cis-regulatory regions in zebrafish. Dev. Dyn. 238, 2409–2417 (2009).

39. Moya, N., Cutts, J., Gaasterland, T., Willert, K. & Brafman, D. A. Endogenous WNT Signaling Regulates hPSC-Derived Neural Progenitor Cell Heterogeneity and Specifies Their Regional Identity. Stem Cell Rep. 3, 1015– 1028 (2014).

40. Manuel, M. et al. The transcription factor Foxg1 regulates the competence of telencephalic cells to adopt subpallial fates in mice. Dev. Camb. Engl. 137, 487–497 (2010).

41. Salehi, H. et al. Differentiation of human ES cell-derived neural progenitors to neuronal cells with regional specific identity by co-culturing of notochord and somite. Stem Cell Res. 8, 120–133 (2012).

42. Salehi, H. et al. Neuronal induction and regional identity by co-culture of adherent human embryonic stem cells with chicken notochords and somites. Int. J. Dev. Biol. 55, 321–326 (2011).

43. Kawauchi, S. et al. Foxg1 promotes olfactory neurogenesis by antagonizing Gdf11. Dev. Camb. Engl. 136, 1453–1464 (2009).

44. Yang, H. et al. SOXE group transcription factors regulates the expression of FoxG1 during inner ear development. Biochem. Biophys. Res. Commun. 623, 96–103 (2022).

45. He, Z. et al. An integrated transcriptomic cell atlas of human neural organoids. Nature 635, 690–698 (2024).

46. Hettige, N. C. et al. FOXG1 dose tunes cell proliferation dynamics in human forebrain progenitor cells. Stem Cell Rep. 17, 475–488 (2022).

47. Mertens, J., Marchetto, M. C., Bardy, C. & Gage, F. H. Evaluating cell reprogramming, differentiation and conversion technologies in neuroscience. Nat. Rev. Neurosci. 17, 424–437 (2016).

48. Wu, C. & Huang, J. Enhancer selectivity across cell types delineates three functionally distinct enhancer-promoter regulation patterns. BMC Genomics 25, 483 (2024).

49. Nott, A. et al. Brain cell type–specific enhancer–promoter interactome maps and disease-risk association. Science 366, 1134–1139 (2019).

50. Paliou, C. et al. Preformed chromatin topology assists transcriptional robustness of Shh during limb development. Proc. Natl. Acad. Sci. 116, 12390–12399 (2019).

51. Andrey, G. et al. Characterization of hundreds of regulatory landscapes in developing limbs reveals two regimes of chromatin folding. Genome Res. 27, 223–233 (2017).

52. Zhang, F. & Lupski, J. R. Non-coding genetic variants in human disease. Hum. Mol. Genet. 24, R102– R110 (2015).

53. Baetens, D., Mendonça, B. b., Verdin, H., Cools, M. & De Baere, E. Non-coding variation in disorders of sex development. Clin. Genet. 91, 163–172 (2017).

54. Liao, J. et al. Deletion of conserved non-coding sequences downstream from NKX2-1: A novel disease-causing mechanism for benign hereditary chorea. Mol. Genet. Genomic Med. 9, e1647 (2021).

55. Benko, S. et al. Disruption of a long distance regulatory region upstream of SOX9 in isolated disorders of sex development. J. Med. Genet. 48, 825–830 (2011).

56. Verdin, H. et al. Microhomology-mediated mechanisms underlie non-recurrent disease-causing microdeletions of the FOXL2 gene or its regulatory domain. PLoS Genet. 9, e1003358 (2013).

57. Kvon, E. Z., Waymack, R., Gad, M. & Wunderlich, Z. Enhancer redundancy in development and disease. Nat. Rev. Genet. 22, 324–336 (2021).

58. Zhang, Y. et al. Rapid Single-Step Induction of Functional Neurons from Human Pluripotent Stem Cells. Neuron 78, 785–798 (2013).

59. Mohajeri, K. et al. Transcriptional and functional consequences of alterations to MEF2C and its topological organization in neuronal models. Am. J. Hum. Genet. 109, 2049 (2022).

60. Langmead, B. & Salzberg, S. L. Fast gapped-read alignment with Bowtie 2. Nat. Methods 9, 357–359 (2012).

61. Raman, L., Dheedene, A., De Smet, M., Van Dorpe, J. & Menten, B. WisecondorX: improved copy number detection for routine shallow whole-genome sequencing. Nucleic Acids Res. 47, 1605–1614 (2019).

62. Sante, T. et al. ViVar: A Comprehensive Platform for the Analysis and Visualization of Structural Genomic Variation. PLOS ONE 9, e113800 (2014).

63. D’haene, B., Vandesompele, J. & Hellemans, J. Accurate and objective copy number profiling using real-time quantitative PCR. Methods 50, 262–270 (2010).

64. D’haene, E. et al. Comparative 3D genome analysis between neural retina and retinal pigment epithelium reveals differential cis-regulatory interactions at retinal disease loci. Genome Biol. 25, 123 (2024).

65. Durand, N. C. et al. Juicer Provides a One-Click System for Analyzing Loop-Resolution Hi-C Experiments. Cell Syst. 3, 95–98 (2016).

66. Li, H. & Durbin, R. Fast and accurate short read alignment with Burrows-Wheeler transform. Bioinformatics 25, 1754–1760 (2009).

67. Open2C, et al. Cooltools: Enabling high-resolution Hi-C analysis in Python. PLOS Comput. Biol. 20, e1012067 (2024).

68. Kruse, K., Hug, C. B. & Vaquerizas, J. M. FAN-C: a feature-rich framework for the analysis and visualisation of chromosome conformation capture data. Genome Biol. 21, 303 (2020).

69. Ewels, P. A. et al. The nf-core framework for community-curated bioinformatics pipelines. Nat. Biotechnol. 38, 276–278 (2020).

70. Meers, M. P., Tenenbaum, D. & Henikoff, S. Peak calling by Sparse Enrichment Analysis for CUT&RUN chromatin profiling. Epigenetics Chromatin 12, 42 (2019).

71. Braun, E. et al. Comprehensive cell atlas of the first-trimester developing human brain. Science 382, eadf1226 (2023).

