## Supplemental Figures for "Non-coding structural variants identify a commonly affected regulatory region steering *FOXG1* transcription in early neurodevelopment"

Hamerlinck L., D'haene E. *et al.*

### SUPPLEMENTARY INFORMATION

**Supplementary Text. Clinical description of new patient included in the manuscript (not included on MedRxiv)**

**Supplementary Figure S1. Identification and finemapping of patient deletion.**
**Supplementary Figure S2. Overview of all CNVs observed in patients overlapping the bigger FOXG1 TAD based on the DECIPHER database.**
**Supplementary Figure S3. Detailed view of the enhancer cluster and TAD boundary.**
**Supplementary Figure S4. Chromatin interaction frequencies at the *FOXG1* and *PRKD1* locus.**
**Supplementary Figure S5. TAD structure conservation at the FOXG1 locus across tissues.**
**Supplementary Figure S6. CTCF binding at the commonly affected regulatory region (CARR) across ENCODE tissues and cell types.**
**Supplementary Figure S7. RAD21 binding at the commonly affected regulatory region (CARR) across ENCODE tissues and cell types.**
**Supplementary Figure S8. Quantification of zebrafish enhancer assay results.**
**Supplementary Figure S9. Single-cell RNA-seq data from human brain development shows high FOXG1 expression in ventral telencephalic NPC and neurons.**
**Supplementary Figure S10. Characterization of NPC populations using regional markers.**
**Supplementary Figure S11. Regulatory dynamics of NPC markers during NPC differentiation.**
**Supplementary Figure S12. Successful differentiation of WT and CRISPR KO models for the EC and TAD boundary towards NPCs.**
**Supplementary Figure S13. Schematic overview of PCR reactions to assess successful generation of the heterozygous KO models.**
**Supplementary Figure S14. Assessing monoclonality and successful editing of the clones within the different genotypes.**

### SUPPLEMENTARY FIGURES

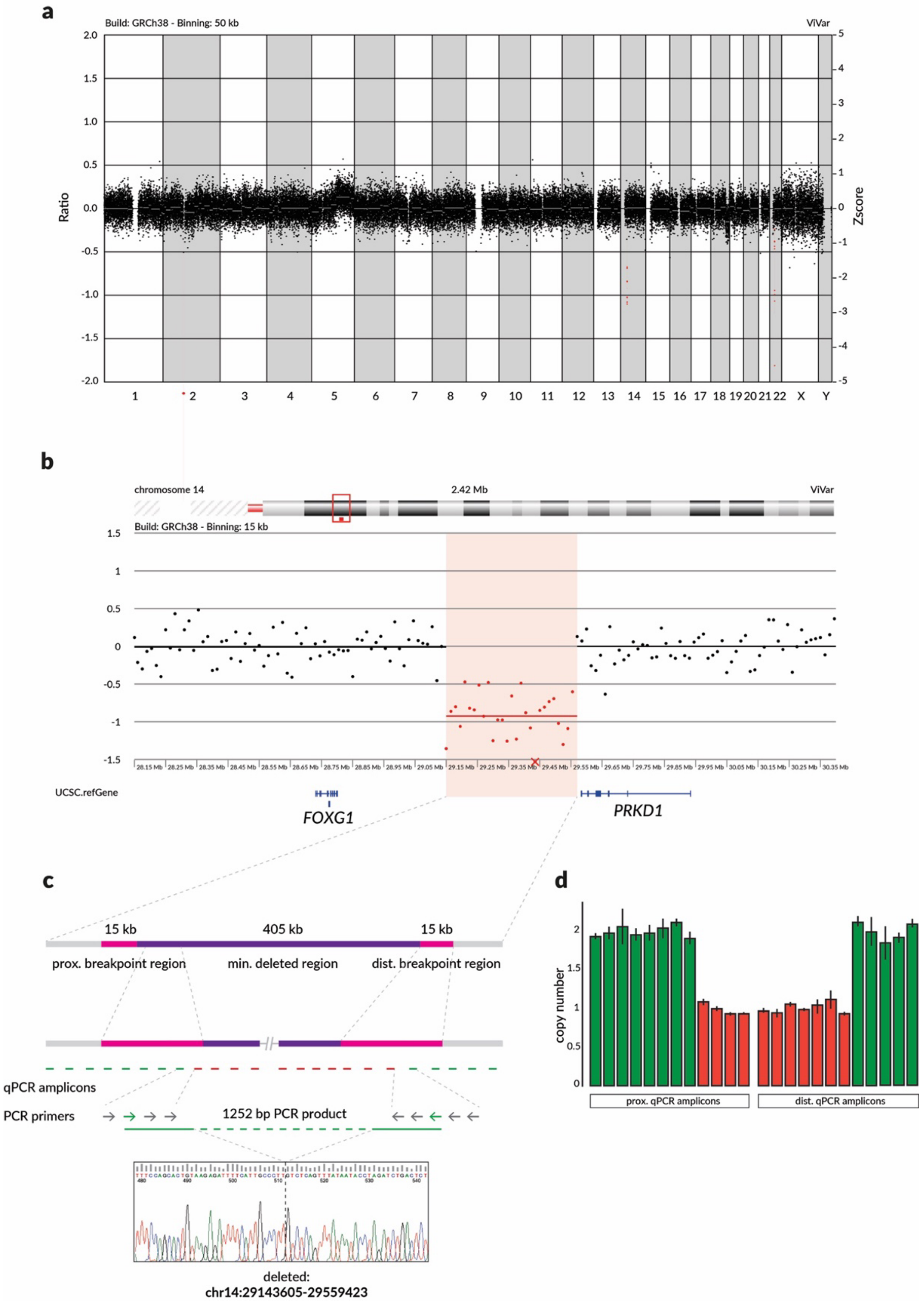


**Figure S1. Identification and finemapping of patient deletion.** (a) Genome-wide view of copy number profile visualized in ViVar and determined through shallow whole genome sequencing on patient lymphoblastoid cells (50 kb binning). (b) Copy number profile showing patient deletion (15 kb binning). (c) Strategy for finemapping of the deletion breakpoints, including design of CNV-qPCR amplicons and PCR primer across the breakpoint region, as well as the Sanger sequencing trace of the PCR product including the breakpoint. (d) Copy number plot for qPCR amplicons designed across the breakpoint regions used to narrow down the deletion breakpoints.

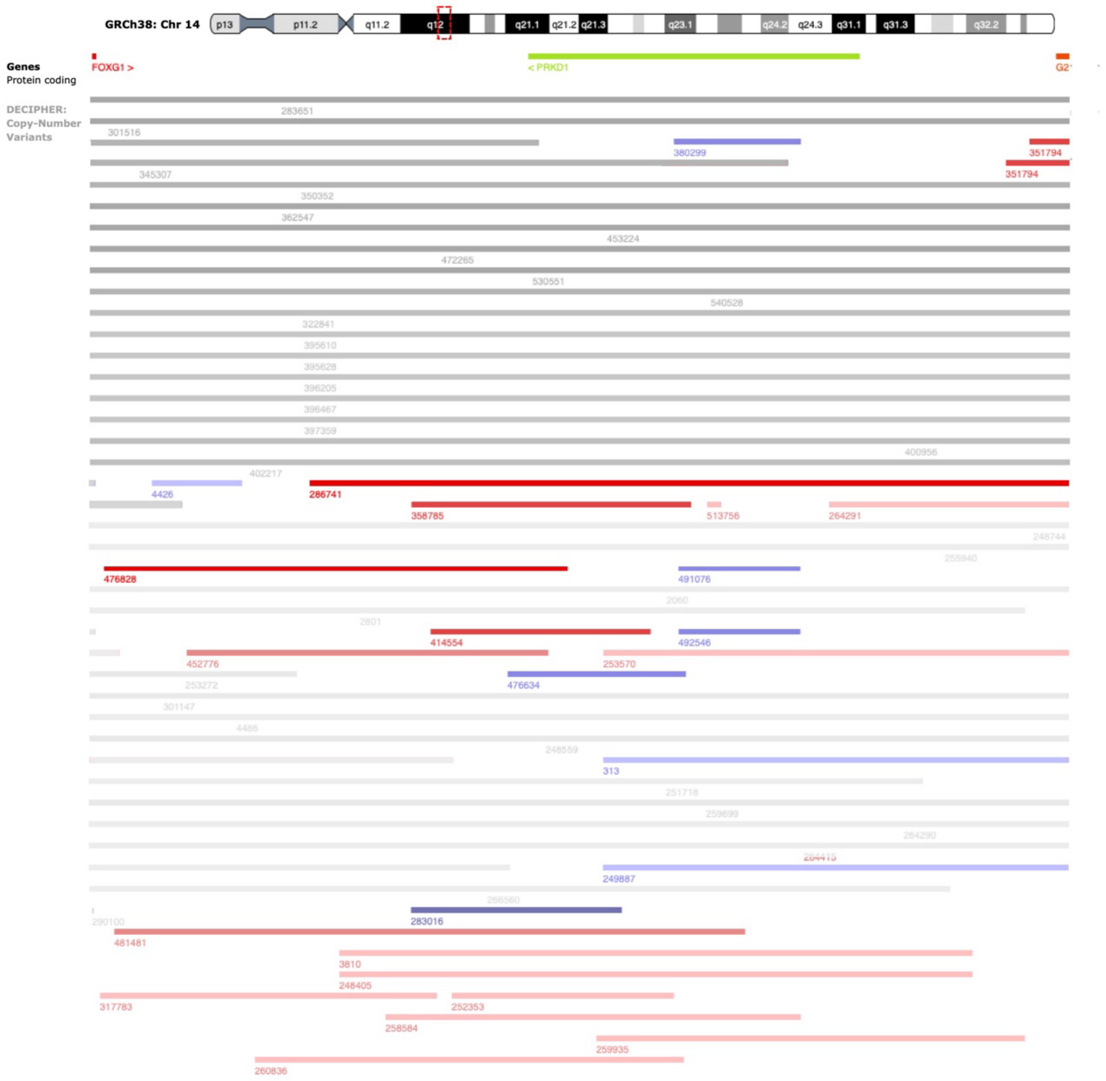


**Figure S2. Overview of all CNVs observed in patients overlapping the bigger FOXG1 TAD based on the DECIPHER database.** Grey: Overlap FOXG1 gene; blue: duplication; red: deletion.

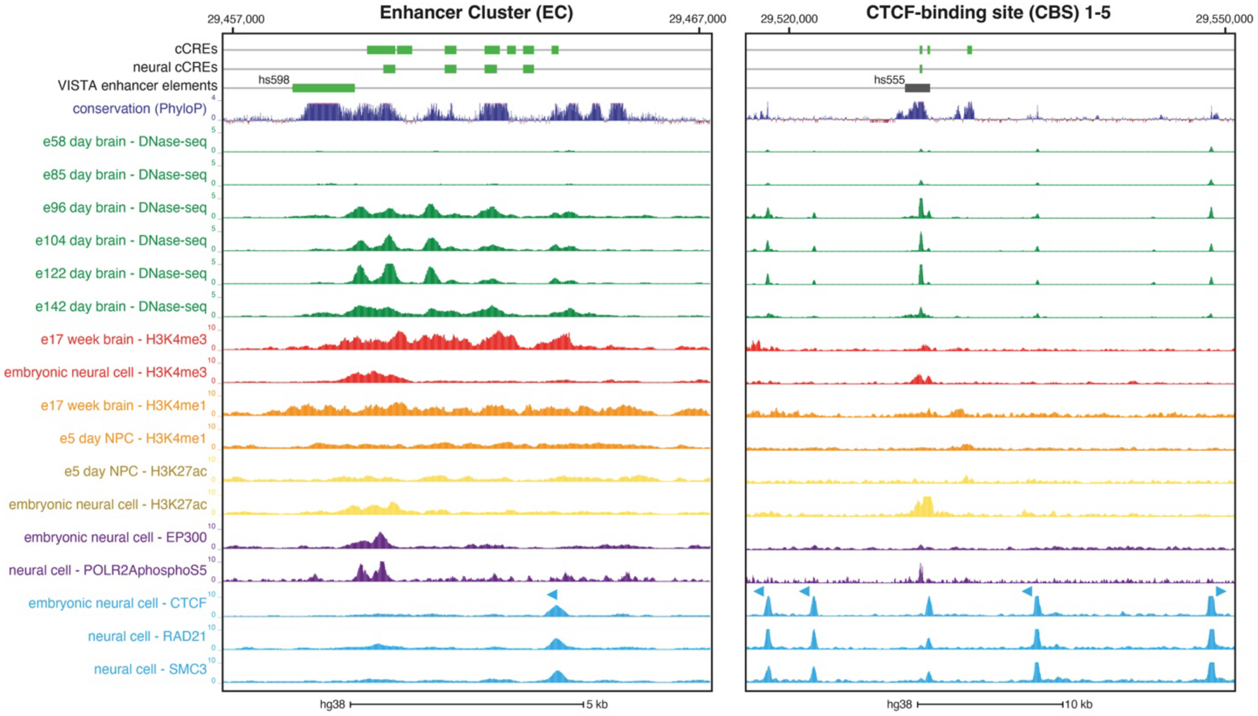


**Figure S3. Detailed view of the enhancer cluster and TAD boundary.** Plot of candidate cis-regulatory elements (cCREs) and publicly available epigenomics data at the enhancer cluster (EC) and TAD boundary region, including: (neural) enhancer-like cCREs based on human DNase I hypersensitive sites (DHSs)[1] and ENCODE SCREEN[2] enhancer-like elements; validated VISTA enhancer elements (positive: green, negative: grey)[3]; PhyloP basewise conservation, accessible chromatin through DNase-seq in embryonic brain, ChIP-seq for regulatory markers H3K4me3, H3K4me1, H3K27ac, as well as active RNA polymerase II (POLR2AphosphoS5) and the P300 cofactor in embryonic brain (embryonic) neural cells and NPCs, ChIP-seq for the architectural protein CTCF and cohesin subunits RAD21 and SMC3 in (embryonic) neural cells.

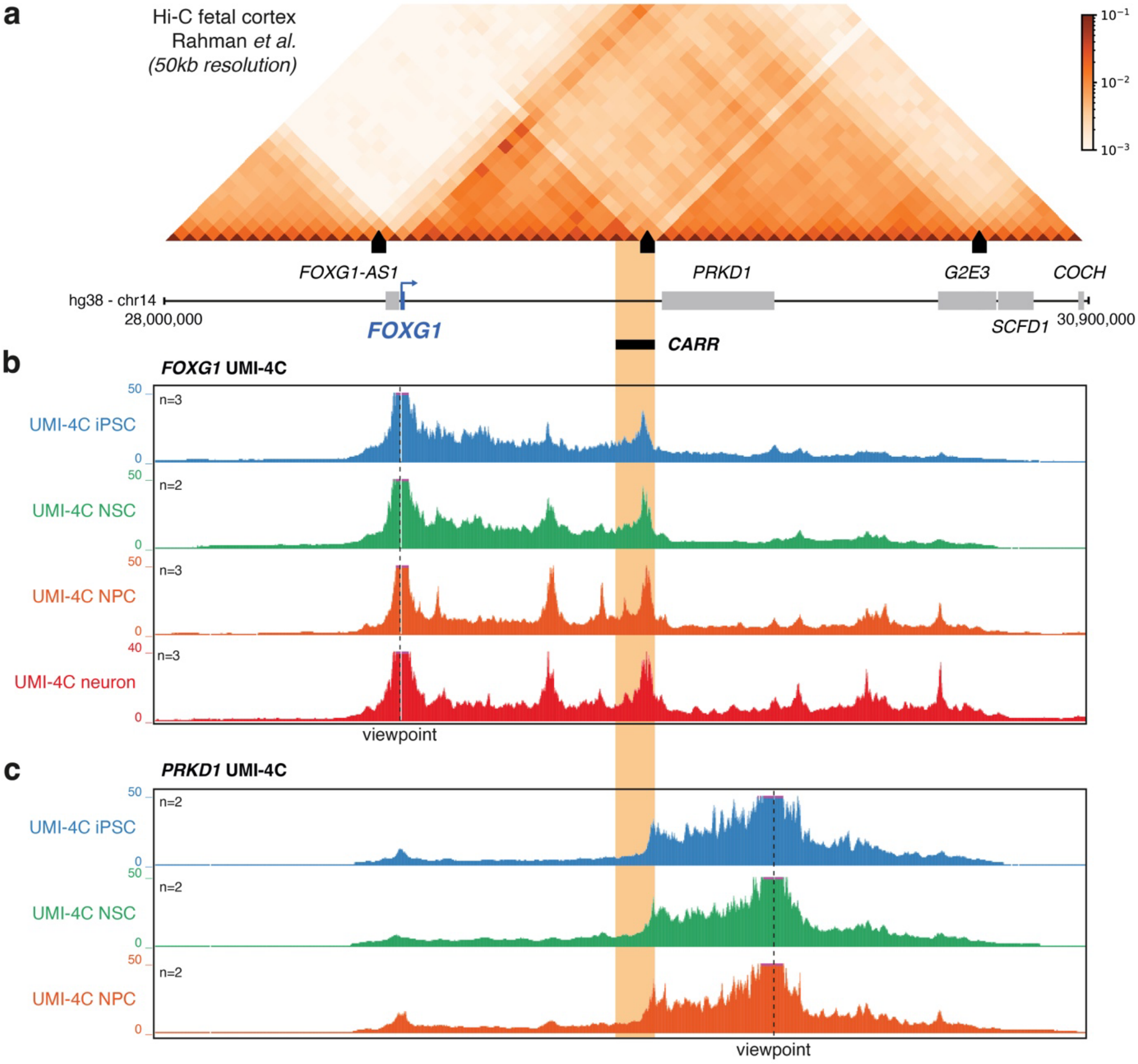


**Figure S4. Chromatin interaction frequencies at the *FOXG1* and *PRKD1* locus.** (a) TAD structure of the *FOXG1* locus as shown by the Hi-C interaction frequency matrix (fetal cortex, 50kb resolution, Rahman *et al.*[4]). TAD boundaries are indicated by black boxes. The commonly affected regulatory region (CARR) is highlighted in orange. (b) UMI-4C interaction frequency profiles (normalized and smoothed) with the *FOXG1* promoter as viewpoint (dashed line) in induced pluripotent stem cells (iPSCs, n=3), neural stem cells (NSCs, n=2), neural progenitor cells (NPCs, n=3) and neurons (n=3). (c) UMI-4C interaction frequency profiles (normalized and smoothed) with the *PRKD1* promoter as viewpoint (dashed line) in induced pluripotent stem cells (iPSCs, n=2), neural stem cells (NSCs, n=2), neural progenitor cells (NPCs, n=2).

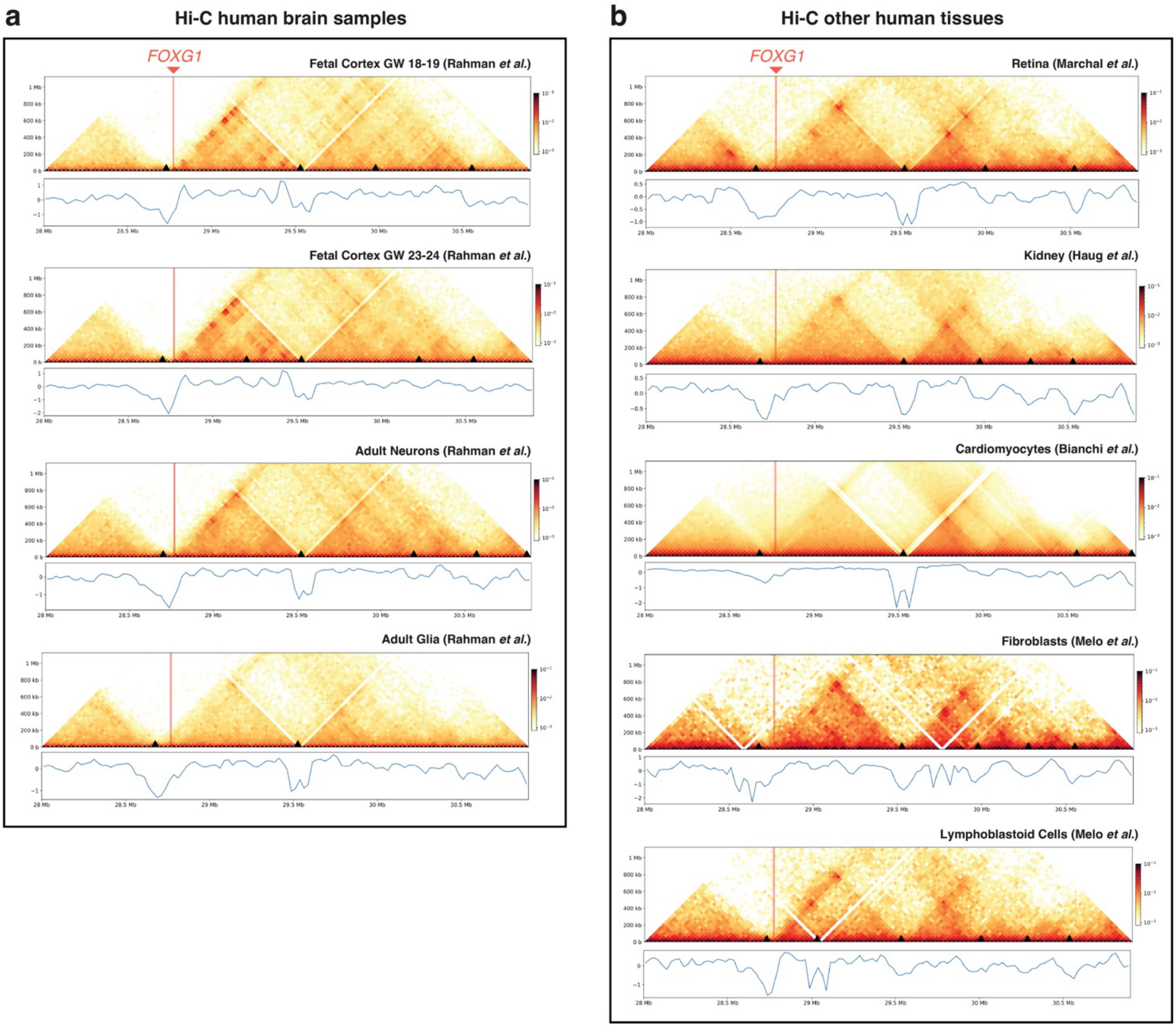


**Figure S5. TAD structure conservation at the *FOXG1* locus across tissues.** Hi-C interaction frequency matrices and diamond insulation score plots (25 kb resolution) depicting *FOXG1* TAD structure in (a) human fetal and adult brain samples[5] and (b) other human tissues[6–9]. TAD boundaries based on insulation score minima are indicated as black boxes.

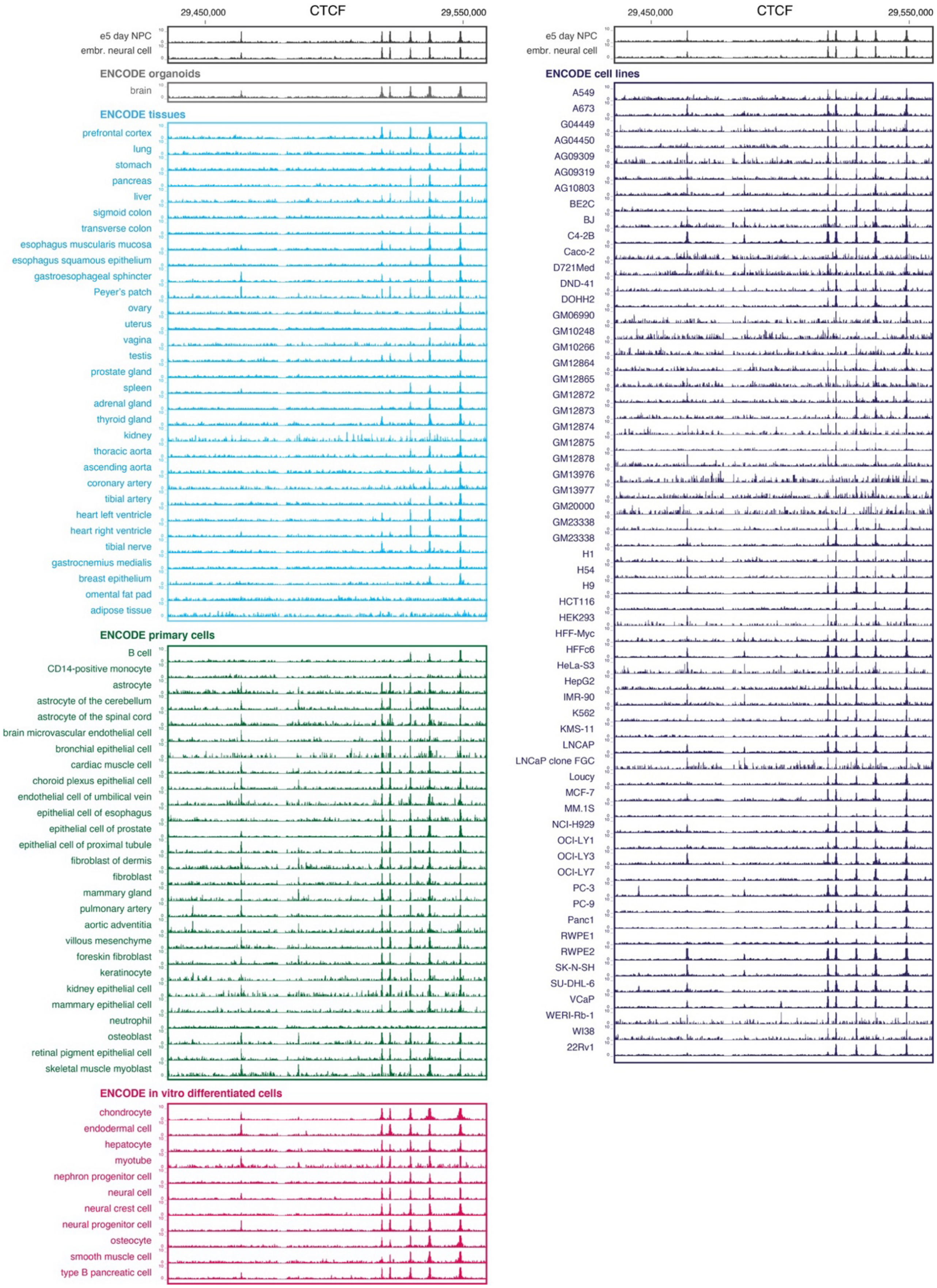


**Figure S6. CTCF binding at the commonly affected regulatory region (CARR) across ENCODE tissues and cell types.**

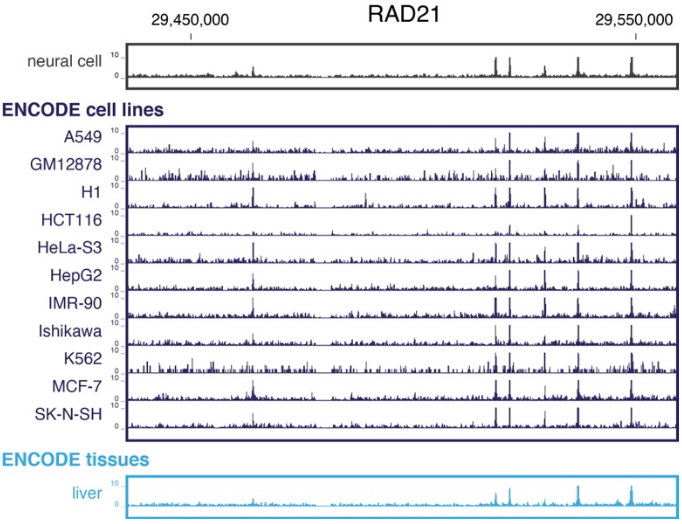


**Figure S7. RAD21 binding at the commonly affected regulatory region (CARR) across ENCODE tissues and cell types.**

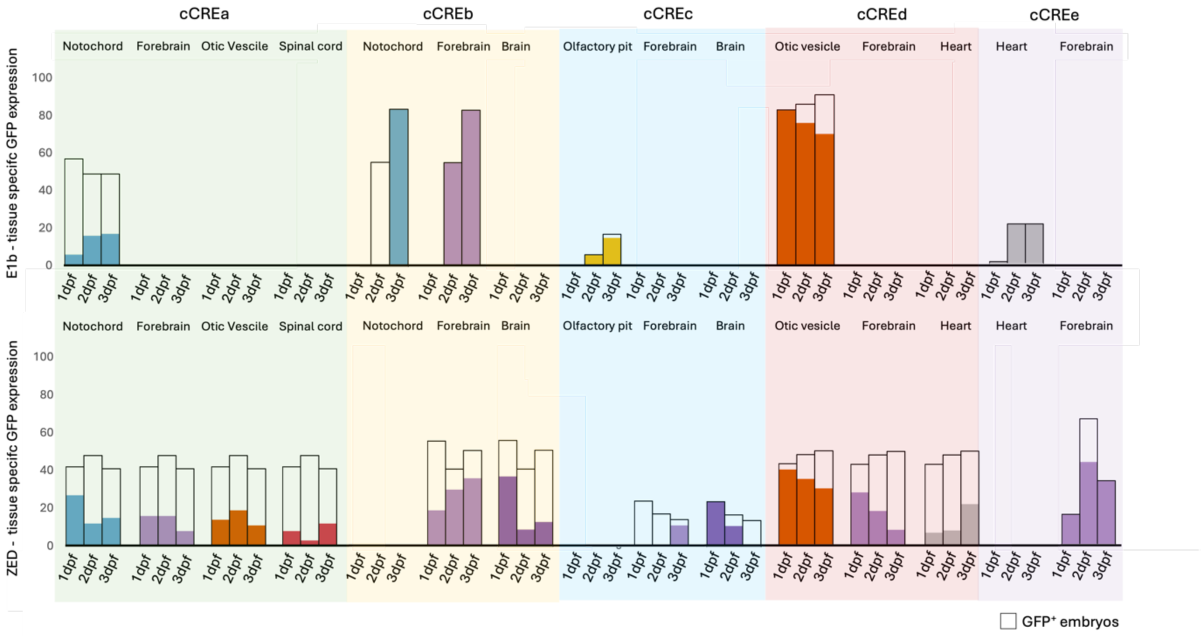


**Figure S8. Quantification of zebrafish enhancer assay results.** Overview of tissue specific GFP expression in zebrafish for our identified enhancer elements: cCREa, cCREb, cCREc, cCREd and cCREe, using either the E1b (up) or ZED (low) vector. The black boxes indicate the number of zebrafish embryos showing general GFP expression after injecting ~100 zebrafish embryos with our construct. The color-coded bars indicate the number of embryos showing tissue specific expression. Blue: notochord; purple: forebrain; orange: otic vesicle; red: spinal cord; grey: heart.

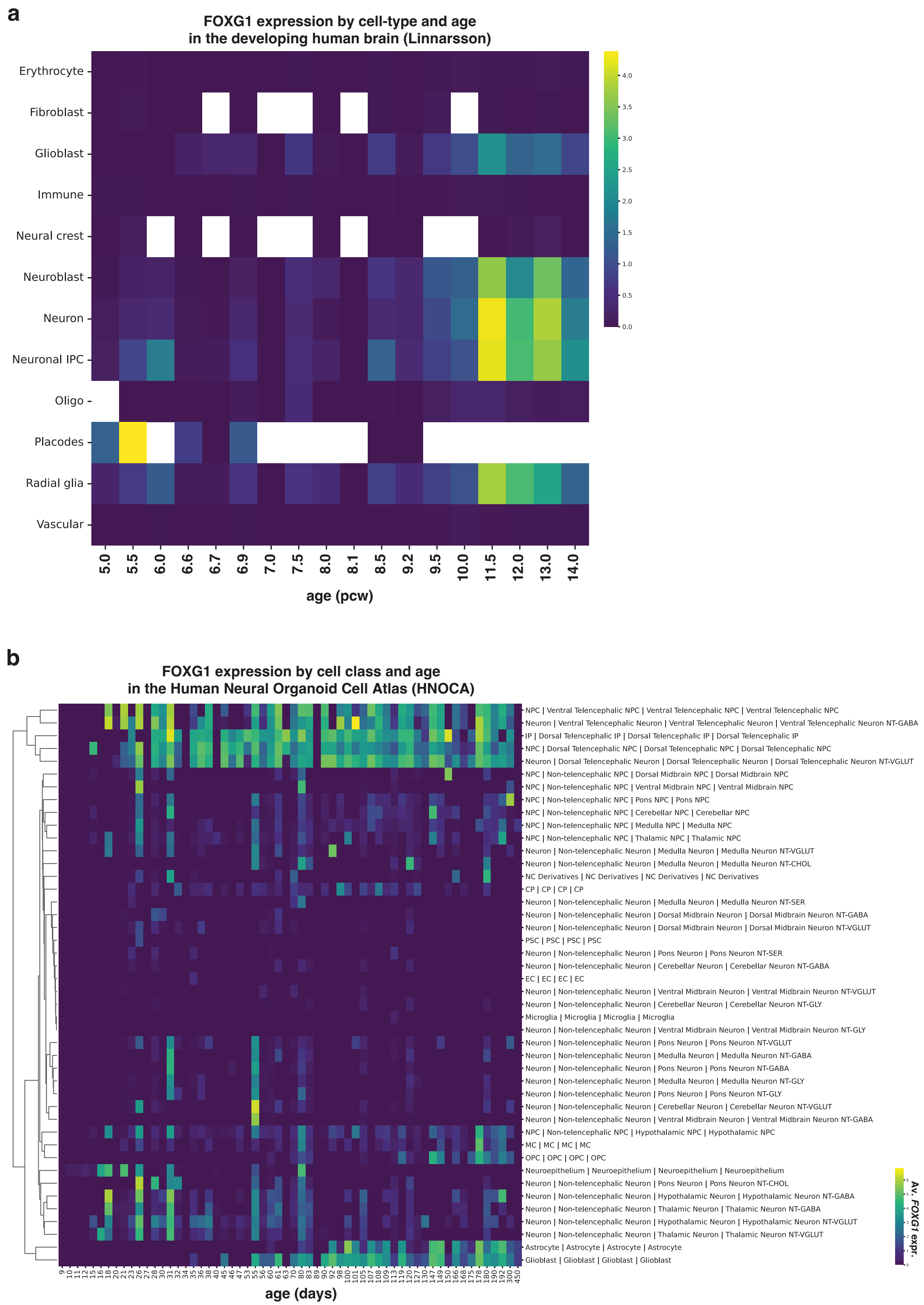


**Figure S9. Single-cell RNA-seq data from human brain development shows high FOXG1 expression in ventral telencephalic NPC and neurons.** (a) FOXG1 mRNA expression per cell-type and age (in post-conceptional weeks, pcw) from single-cell RNA-seq data by Braun et al.[10] (b) FOXG1 mRNA expression per cell class and differentiation stage from the Human Neural Organoid Cell Atlas (HNOCA)[11].

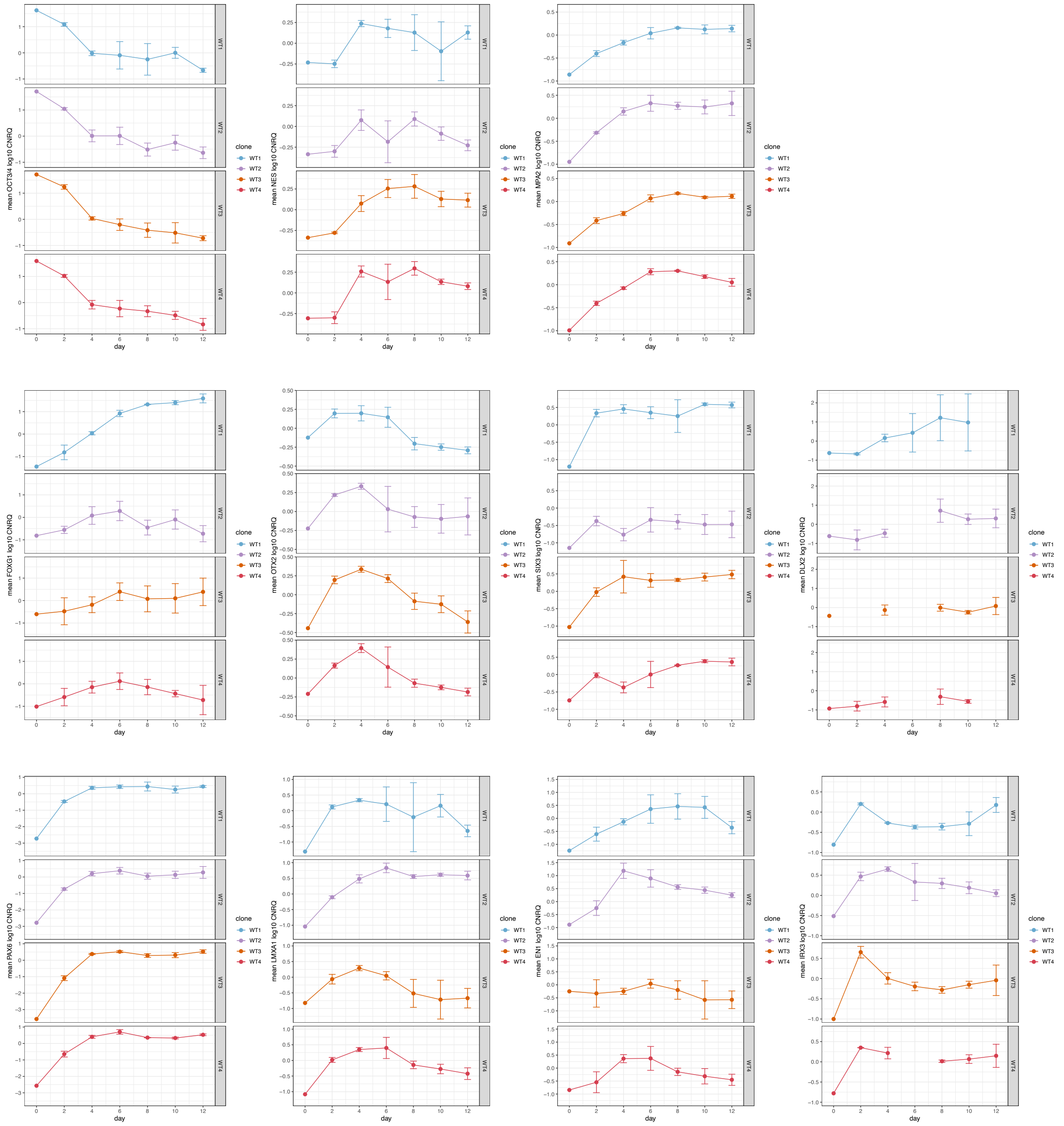


**Figure S10. Characterization of NPC populations using regional markers.** RNA expression pluripotency markers (OCT3/4), neural markers (NES, MAP2) and various NPC markers for different brain regions: FOXG1, OTX2, SIX3 and DLX2 (forebrain); PAX6 (forebrain/midbrain); LMX1A (midbrain); EN1 and IRX3 (midbrain/hindbrain) during a 12-day NPC differentiation for 4 biological replicates.

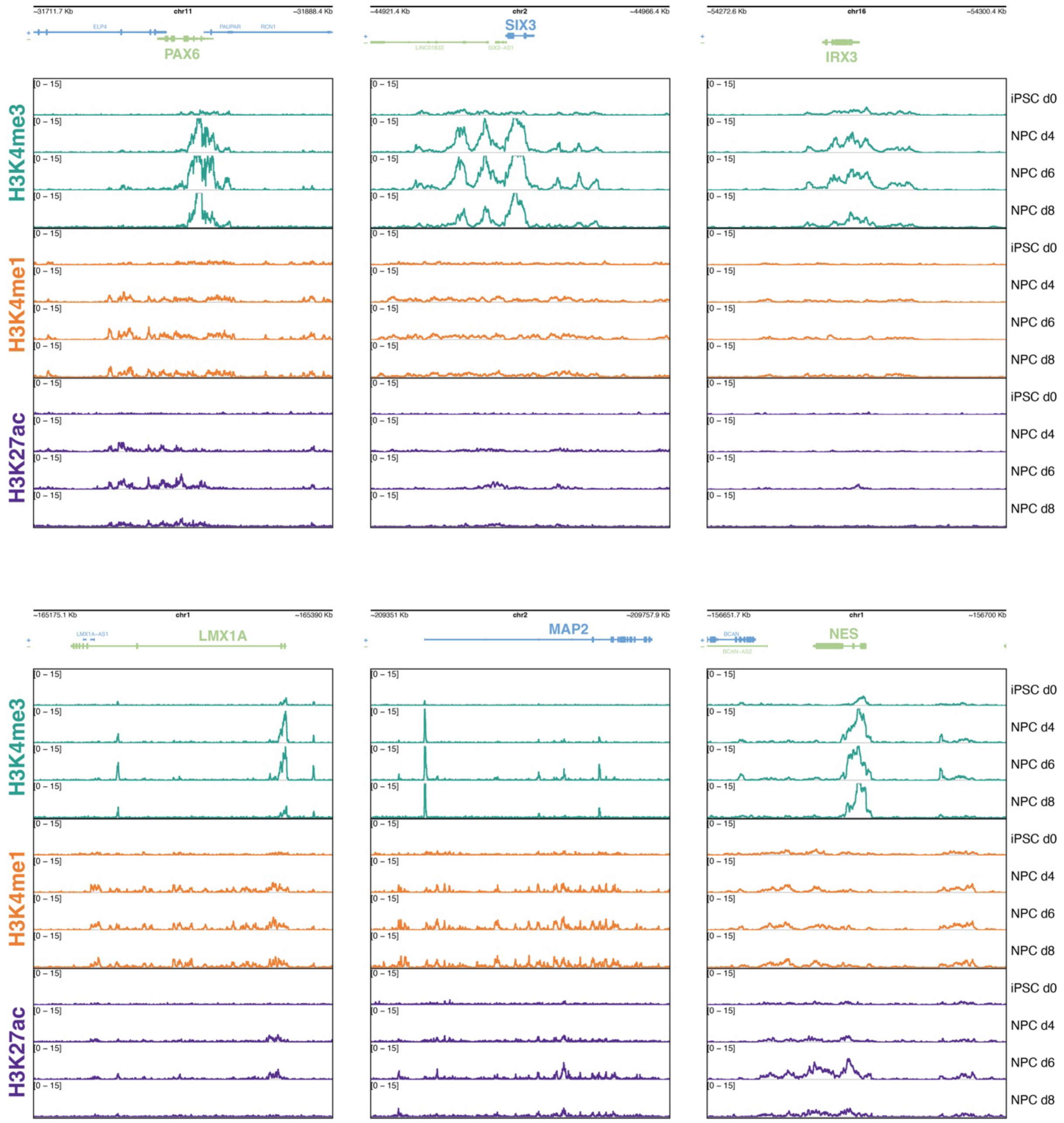


**Figure S11. Regulatory dynamics of NPC markers during NPC differentiation.** CUT&RUN signal tracks for H3K4me3 (active promotor marker), H3K4me1 and H3K27ac (active enhancer markers) and CTCF (TAD) for various NPC markers for different brain regions: PAX6 (forebrain/midbrain); SIX3 (forebrain); IRX3 (midbrain/hindbrain); LMX1A (midbrain); MAP2 (neural); NES (pan-neural marker) during an 8-day neural progenitor cell (NPC) differentiation from induced pluripotent stem cells (iPSC) (n=2).

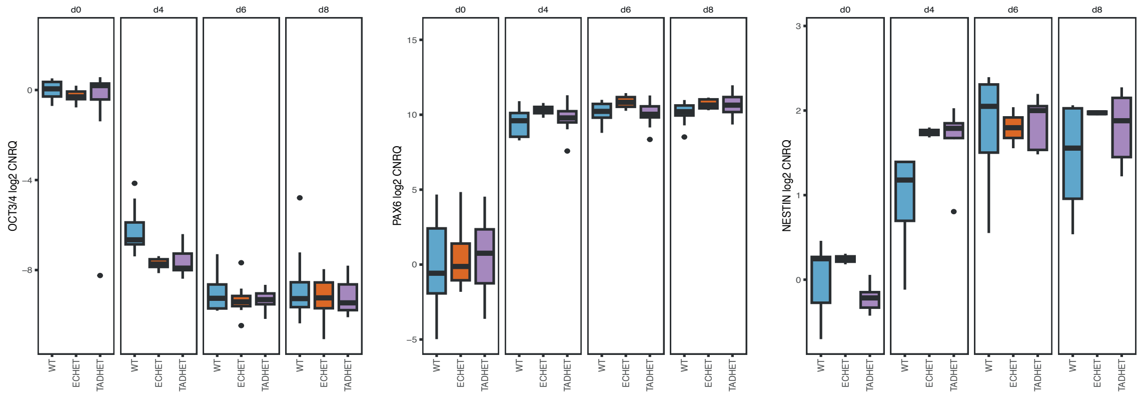


**Figure S12. Successful differentiation of WT and CRISPR KO models for the EC and TAD boundary towards NPCs.** RNA expression of OCT3/4 (iPSC marker) and PAX6 (NPC marker) and NES (neural marker) during an 8-day NPC differentiation for 3 genotypes: wildtype (WT), heterozygous deletion of the enhancer cluster (ECHET) and heterozygous deletion of the TAD boundary (TADHET). Each genotype encompasses at least 3 biological replicates (WT n=6; ECHET n=3; TADHET n=6). RNA expression data is visualized in log2, scaled to iPSCs.

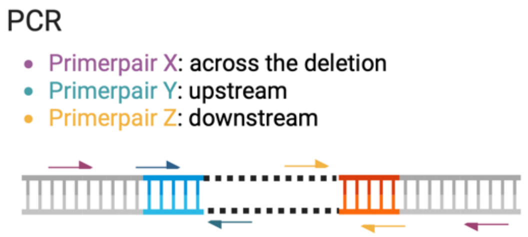


**Figure S13. Schematic overview of PCR reactions to assess successful generation of the heterozygous KO models.**

## EC_QE1_A6

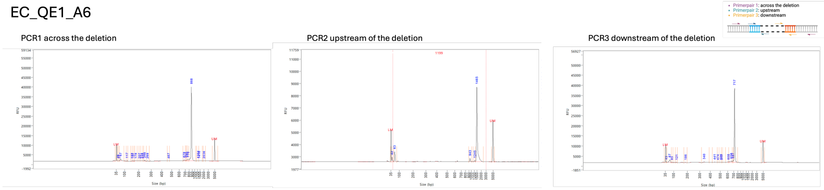


CNV-sequencing 5kb bin

| | |
|---|---|
| EC_gRNA_up_ext_1 | 29456780 |
| EC_gRNA_down | 29467257 |

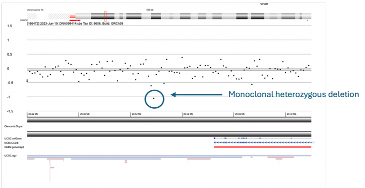


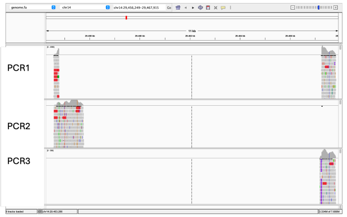


NOT MONOCLONAL

## EC_QE1_A12

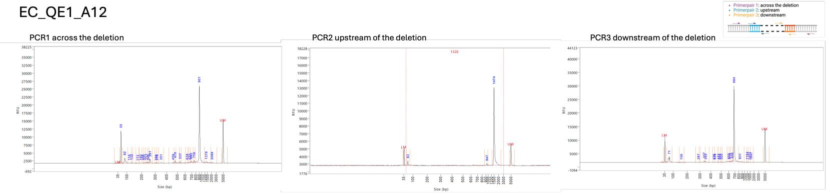


CNV-sequencing 5kb bin

| | |
|---|---|
| EC_gRNA_up_ext_1 | 29456780 |
| EC_gRNA_down | 29467257 |

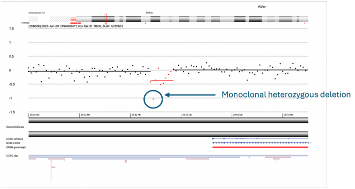


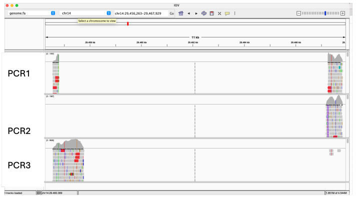


## EC_QE2_A4

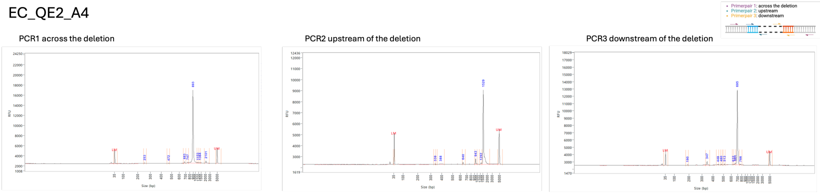


CNV-sequencing 5kb bin

| | |
|---|---|
| EC_gRNA_up_ext_1 | 29456780 |
| EC_gRNA_down | 29467257 |

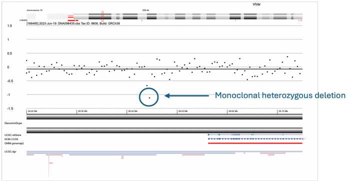


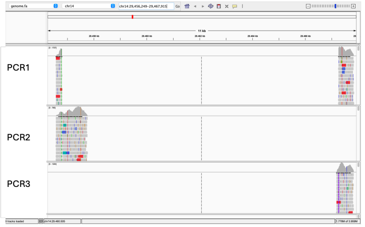


## EC_QE1_A4

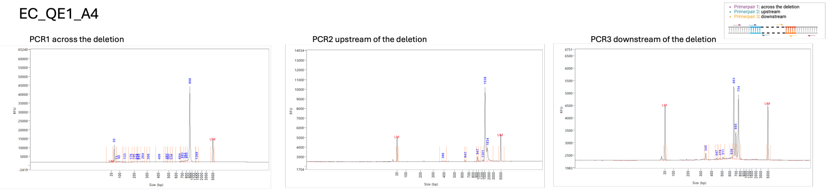


CNV-sequencing 5kb bin

| | |
|---|---|
| EC_gRNA_up_ext_1 | 29456780 |
| EC_gRNA_down | 29467257 |

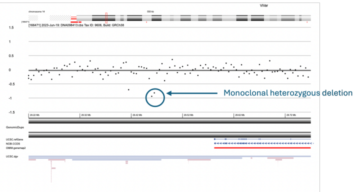


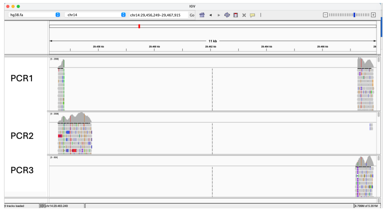


NOT MONOCLONAL

## EC_QE2_A7

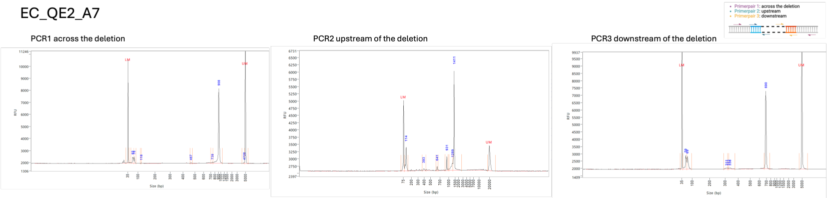


CNV-sequencing 5kb bin

| | |
|---|---|
| EC_gRNA_up_ext_1 | 29456780 |
| EC_gRNA_down | 29467257 |

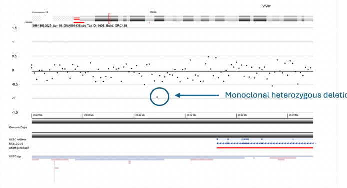


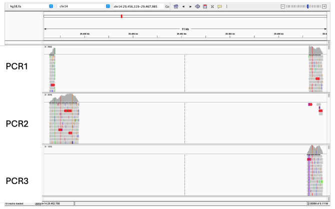


NOT MONOCLONAL

## EC_QE1_B6

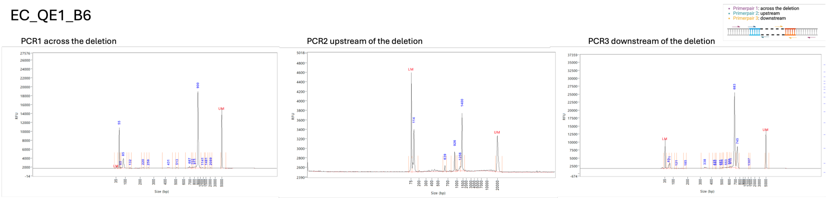


CNV-sequencing 5kb bin

| | |
|---|---|
| EC_gRNA_up_ext_1 | 29456780 |
| EC_gRNA_down | 29467257 |

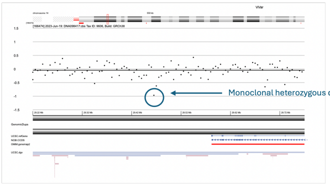


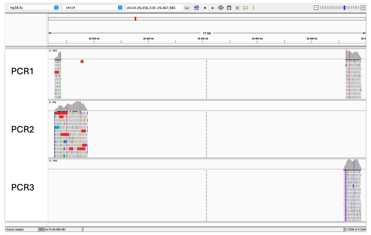

### TAD_QE1_B2

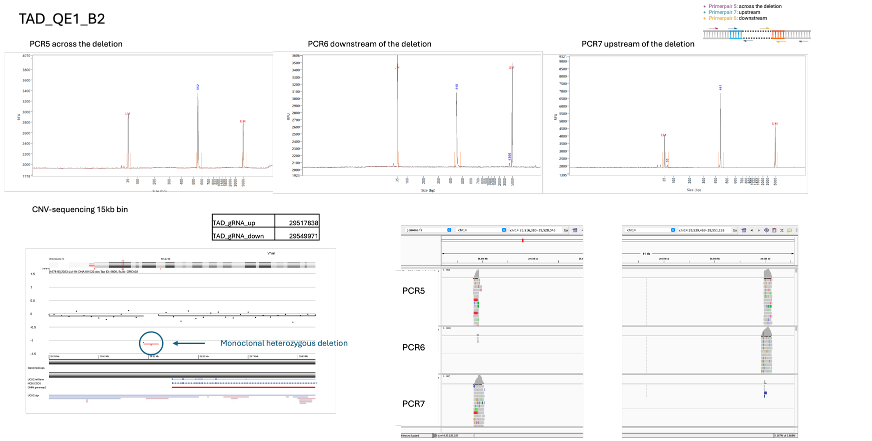


| | |
|---|---|
| TAD_gRNA_up | 29517838 |
| TAD_gRNA_down | 29549971 |

### TAD_B4

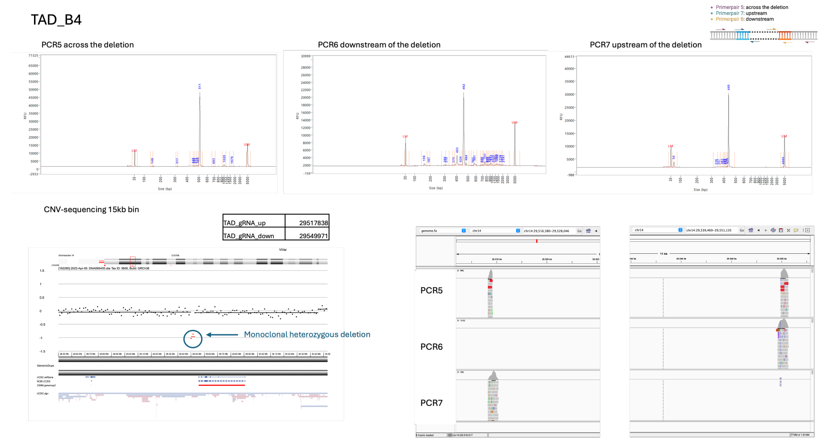


| | |
|---|---|
| TAD_gRNA_up | 29517838 |
| TAD_gRNA_down | 29549971 |

### TAD_H11

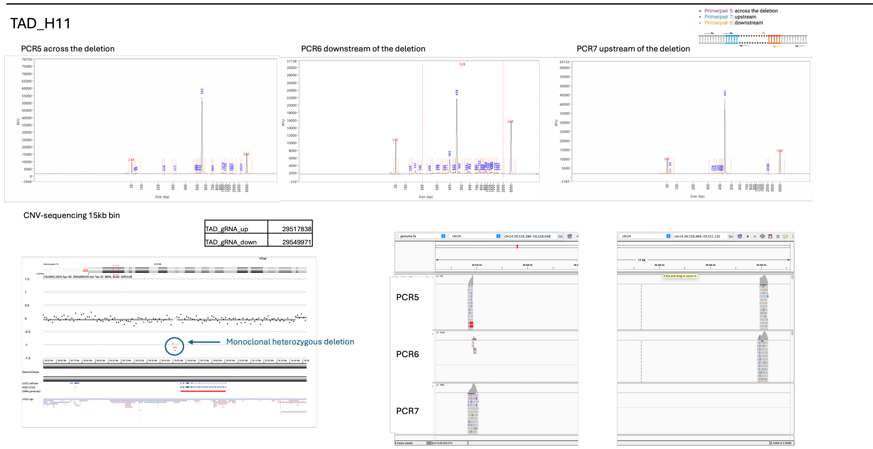


| | |
|---|---|
| TAD_gRNA_up | 29517838 |
| TAD_gRNA_down | 29549971 |

### TAD_QE1_B4

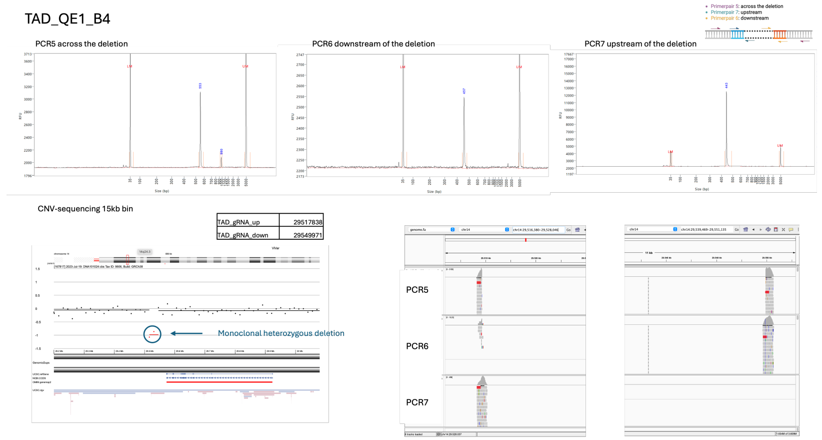


| | |
|---|---|
| TAD_gRNA_up | 29517838 |
| TAD_gRNA_down | 29549971 |

### TAD_QE1_F5

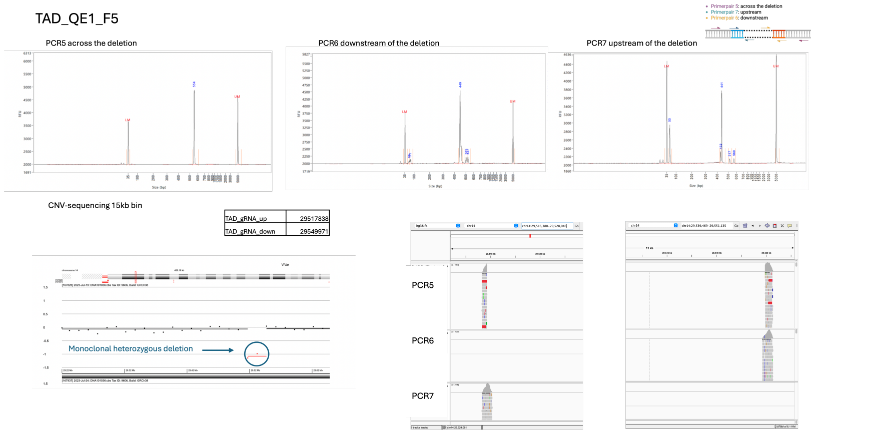


| | |
|---|---|
| TAD_gRNA_up | 29517838 |
| TAD_gRNA_down | 29549971 |

### TAD_QE1_E9

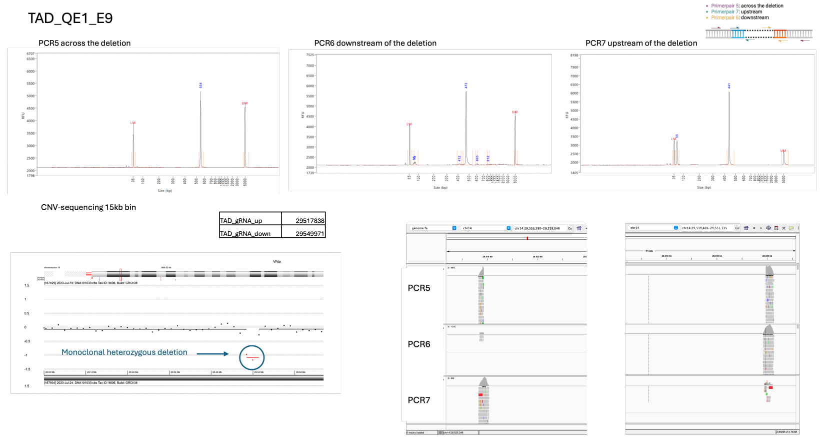


| | |
|---|---|
| TAD_gRNA_up | 29517838 |
| TAD_gRNA_down | 29549971 |

## WT_EC_QE1_F7

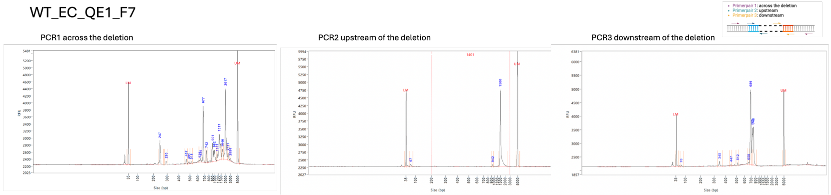


CNV-sequencing 5kb bin

| | |
|---|---|
| EC_gRNA_up_ext_1 | 29456780 |
| EC_gRNA_down | 29467257 |

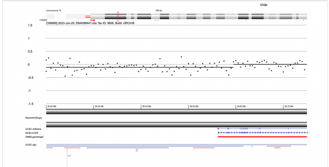

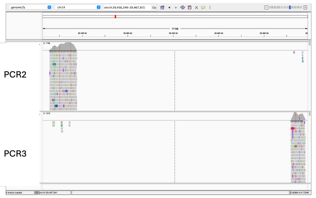


## WT_EC_QE1_E3

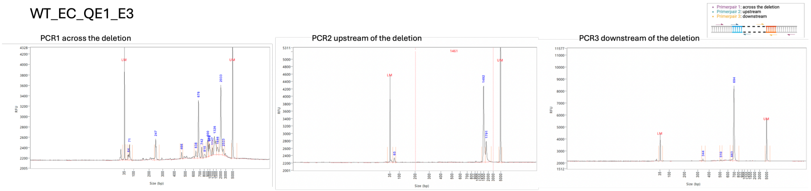


CNV-sequencing 5kb bin

| | |
|---|---|
| EC_gRNA_up_ext_1 | 29456780 |
| EC_gRNA_down | 29467257 |

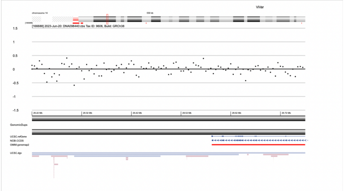

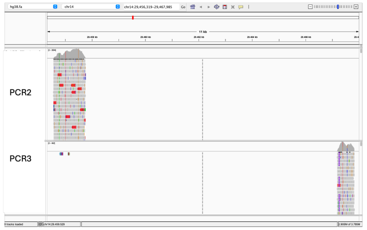


### WT_TAD_C1

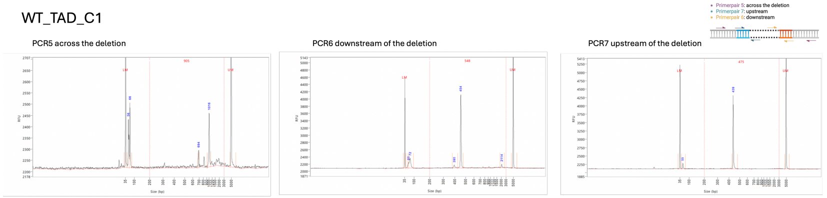


CNV-sequencing 15kb bin

| | |
|---|---|
| TAD_gRNA_up | 29517838 |
| TAD_gRNA_down | 29549971 |

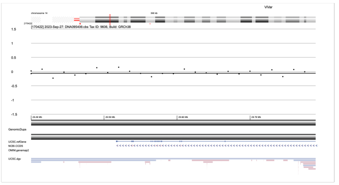

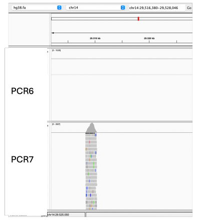


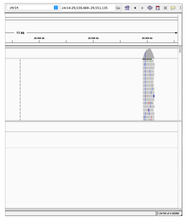

### WT_TAD_H12

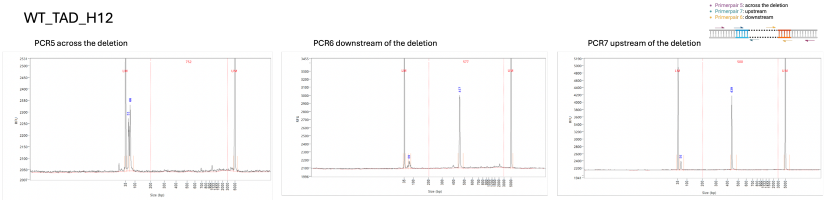


CNV-sequencing 15kb bin

| | |
|---|---|
| TAD_gRNA_up | 29517838 |
| TAD_gRNA_down | 29549971 |

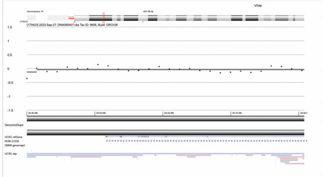

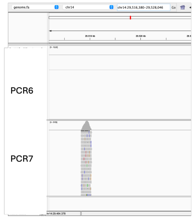


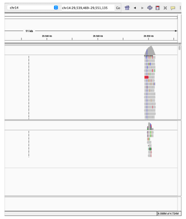

## WT_AF_E8

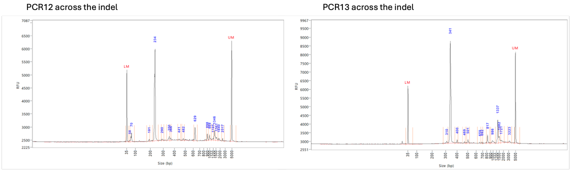


CNV-sequencing 50kb bin

Variants

qDNAseq

| exp | chr | | start | end | size | ratio | type | | |
|---|---|---|---|---|---|---|---|---|---|
| 162259 | 2 | 2p13.1p13.1 | 73.6 | 73.7 | 0.1 | -0.36 | del | ISCN | 1 dgvStudy, |
| 162259 | 9 | 9p23p23 | 12 | 12.1 | 0.1 | -1.05 | del | ISCN | 38 dgvStudies, |
| 162259 | 9 | 9q33.1q33.1 | 119.05 | 119.15 | 0.1 | 0.7 | tripl | ISCN | 1 dgvStudy, |
| 162259 | 14 | 14q11.2q11.2 | 18.2 | 20 | 1.8 | -0.37 | del | ISCN | |
| 162259 | X | 23q28null | 155.9 | 156.05 | 0.15 | 0.52 | dup | ISCN | |

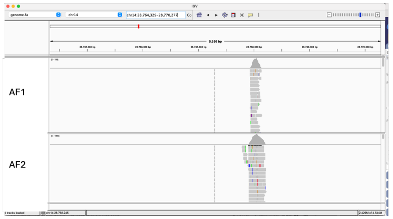

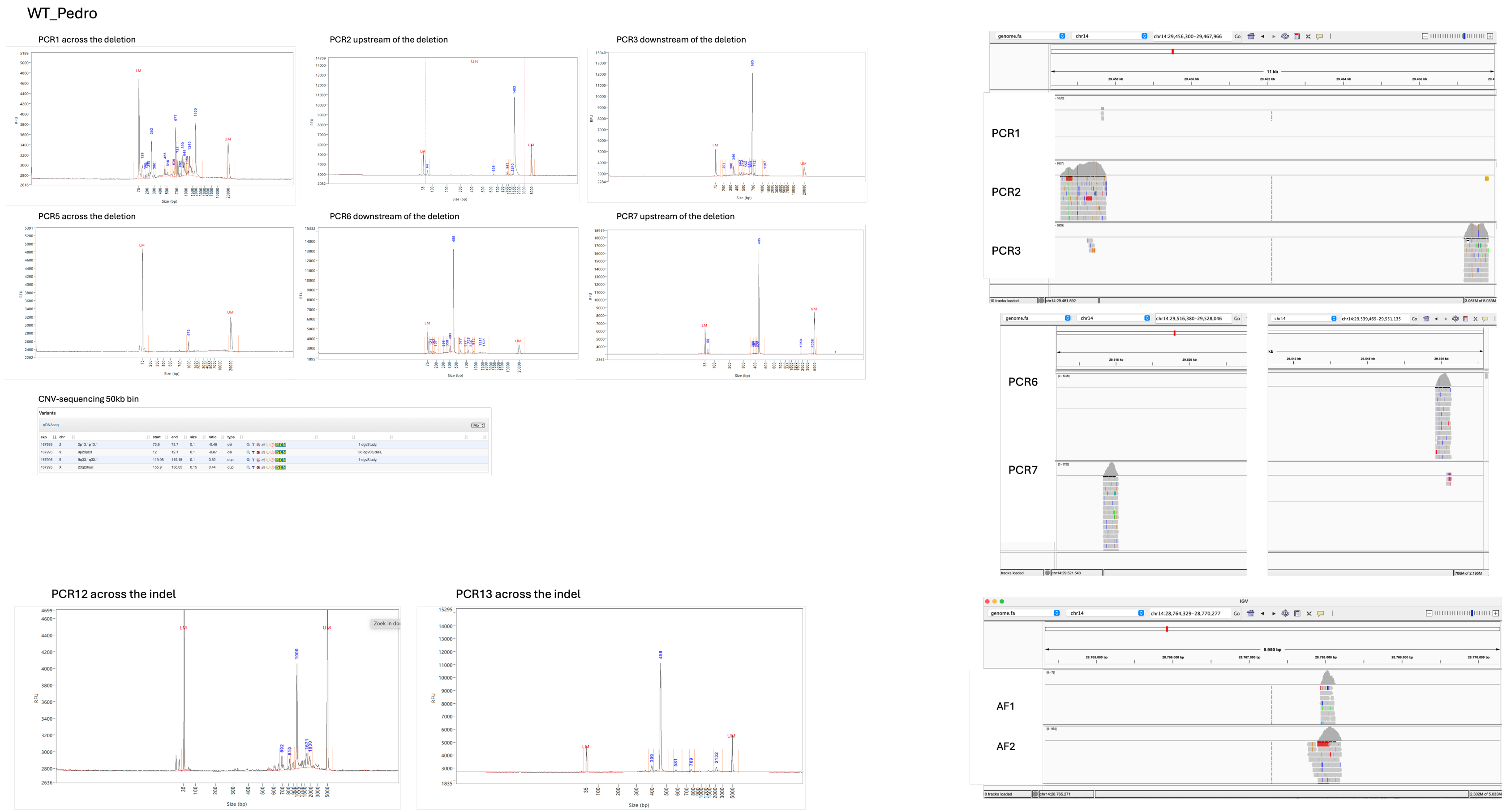


**Figure S14. Assessing monoclonality and successful editing of the clones within the different genotypes.** Successful editing is assessed via DNA isolation and amplification of the target region, followed by targeted next-generation-sequencing (NGS). For heterozygous deletions, a PCR fragment is expected in all three PCR reactions. Finally, monoclonality is confirmed via DNA isolation and amplification of the target region, followed by targeted NGS (indels) and CNV-seq (kb-sized deletions).

### REFERENCES


1. Meuleman, W. *et al.* Index and biological spectrum of human DNase I hypersensitive sites. *Nature* **584**, 244–251 (2020).
2. Moore, J. E. *et al.* Expanded encyclopaedias of DNA elements in the human and mouse genomes. *Nature* **583**, 699–710 (2020).
3. Visel, A., Minovitsky, S., Dubchak, I. & Pennacchio, L. A. VISTA Enhancer Browser--a database of tissue-specific human enhancers. *Nucleic Acids Res.* **35**, D88-92 (2007).
4. Rahman, S. *et al.* Lineage specific 3D genome structure in the adult human brain and neurodevelopmental changes in the chromatin interactome. *Nucleic Acids Res.* **51**, 11142–11161 (2023).
5. Rahman, S. *et al.* Lineage specific 3D genome structure in the adult human brain and neurodevelopmental changes in the chromatin interactome. *Nucleic Acids Res.* **51**, 11142–11161 (2023).
6. Marchal, C. *et al.* High-resolution genome topology of human retina uncovers super enhancer-promoter interactions at tissue-specific and multifactorial disease loci. *Nat. Commun.* **13**, 5827 (2022).
7. Bianchi, V. *et al.* Detailed Regulatory Interaction Map of the Human Heart Facilitates Gene Discovery for Cardiovascular Disease. 705715 Preprint at https://doi.org/10.1101/705715 (2019).
8. Haug, S. *et al.* Multi-omic analysis of human kidney tissue identified medulla-specific gene expression patterns. *Kidney Int.* **105**, 293–311 (2024).
9. Melo, U. S. *et al.* Hi-C Identifies Complex Genomic Rearrangements and TAD-Shuffling in Developmental Diseases. *Am. J. Hum. Genet.* **106**, 872–884 (2020).
10. Braun, E. *et al.* Comprehensive cell atlas of the first-trimester developing human brain. *Science* **382**, eadf1226 (2023).
11. He, Z. *et al.* An integrated transcriptomic cell atlas of human neural organoids. *Nature* **635**, 690–698 (2024).
